# Co-prescription of Metformin and Antipsychotics in Severe Mental Illness: A UK Primary Care Cohort Study

**DOI:** 10.1101/2024.12.03.24318407

**Authors:** Luiza Farache Trajano, Joseph F. Hayes, Naomi Launders, Neil M. Davies, David P. J. Osborn, Alvin Richards-Belle

## Abstract

**Background:** Metformin is a pharmacological candidate to mitigate second-generation antipsychotic (SGA)-induced weight gain in patients with severe mental illnesses (SMI).

**Objective:** To evaluate the incidence, prevalence, and demographic patterns of metformin co-prescription among patients diagnosed with SMI initiating SGAs. To estimate the impact of co-prescription on weight.

**Methods:** A cohort study of patients diagnosed with SMI initiating aripiprazole, olanzapine, quetiapine, or risperidone in 2005-2019 using primary care data from Clinical Practice Research Datalink. We estimated cumulative incidence and period prevalences of co-prescription and explored prescribing differences by demographic and clinical factors. We compared weight change among patients prescribed an SGA only versus an SGA plus metformin, accounting for confounders using linear regression.

**Findings:** Among 26,537 patients initiating SGAs, 4652 were ever prescribed metformin and 21,885 were not. Two-year incidence of first metformin prescription was 3.3%. The SGA plus metformin group were more ethnically diverse, had greater social deprivation, more comorbidities, and higher baseline weight (mean 90.4 vs. 76.8 kg). By two years post-SGA initiation, mean weight in the SGA- only group had changed by +4.16% (95% CI, -1.26 to +9.58) compared to -0.65% (95% CI, -4.26 to +2.96) in the SGA plus metformin group. After confounder adjustment, the two-year mean difference in weight with metformin co-prescription was -1.48 kg (95% CI, -4.03 to 1.07) among females and -1.84 kg (95% CI, -4.67 to 0.98) among males.

**Conclusion:** Metformin is infrequently co-prescribed, despite established efficacy and guidelines.

**Clinical implications:** Primary and secondary care collaboration should be strengthened and barriers to co-prescribing addressed.

## BACKGROUND

Second-generation antipsychotic (SGA) medications are effective in the treatment of severe mental illness (SMI), including schizophrenia and bipolar disorder[1], [2], [3]. In the UK, four SGAs - olanzapine, quetiapine, risperidone and aripiprazole - account for 79% of all primary care antipsychotic prescriptions for patients diagnosed with SMI[4]. These SGAs can have a significant side effect profile, including weight gain; particularly olanzapine, quetiapine, risperidone[5], [6]. Although aripiprazole is generally associated with lower metabolic risk, some individuals experience significant weight gain[7]. During the first three years following SGA initiation, 80% of patients with first-episode psychosis gain a clinically-significant amount of weight (≥7% of their baseline body weight) [8]. Antipsychotic-induced weight gain (AIWG) leads to an increased risk of cardiovascular and metabolic conditions including myocardial infarction, cerebrovascular accidents, dyslipidaemia, and diabetes [5], [9], [10]. Moreover, AIWG can lead to psychological distress, non-adherence, and early antipsychotic discontinuation[8], [11].

Non-pharmacological approaches, such as cognitive and behavioural interventions, physical activity, and dietary modifications, are effective at managing AIWG for some, but not all, patients [12], [13]. Randomised clinical trials (RCTs) investigating the efficacy of these non-pharmacological interventions have suffered high attrition, indicative of adherence challenges[14]. Metformin hydrochloride (metformin), a glucose-lowering medication, licensed for managing type 1 and type 2 diabetes mellitus and polycystic ovarian syndrome (PCOS) [15], has demonstrated significant efficacy in the management of AIWG[16], [17], [18], [19], out-performing other pharmacological candidates in RCTs and meta-analyses[20], [21]. In 2016, the British Association for Psychopharmacology (BAP) recommended adjunctive metformin to manage AIWG and reduce diabetes risk[22]. A 2023 National Institute of Health and Care Research (NIHR) call for research into the clinical and cost- effectiveness of the metformin in preventing AIWG in first-episode psychosis demonstrated a need for further research in this area [23]. In 2024, National Institute for Health and Care Excellence (NICE) reviewed 18 studies and concluded that metformin consistently reduces AIWG by approximately 3 kg when compared to placebo[24]. NICE’s review recommended that their guidelines on psychosis and schizophrenia in adults[25] are updated to ‘explore the role of metformin for managing AIWG’, including off-label use[24]. Despite evidence on efficacy and guideline recommendations, the extent to which metformin is currently being used to manage AIWG in UK primary care is unclear. Understanding current practice will help to inform the implementation of guideline recommendations and highlight unmet need.

### Aim and objectives

In this study, we aimed to described patterns of co-prescribing of SGAs and metformin for people diagnosed with SMI in UK primary care between 2005 until 2019. Our specific objectives were as follows:

1. To describe the cumulative incidence and prevalence of metformin co-prescription in patients diagnosed with SMI newly prescribed aripiprazole, olanzapine, quetiapine, or risperidone in UK primary care.
2. To describe the co-prescription of metformin and SGAs according to demographic and clinical factors and explore if these factors differ by potential metformin indication.
3. To describe changes in weight up to two-years post-SGA initiation among patients co- prescribed metformin plus an SGAs versus patients prescribed an SGA only.

## METHODS

### Study design

We conducted an observational longitudinal cohort study to investigate SGA and metformin co-prescribing in UK primary care from January 2005 to December 2019.

### Data source

We used data from the Clinical Practice Research Datalink (CPRD), which includes two anonymised databases: GOLD[26] and Aurum[27]. These databases include current and historical primary care records for more than 62 million patients [28]. We used the April 2023 and May 2022 database builds of GOLD and Aurum, respectively. For patients registered at primary care practices in England, CPRD data are linked to hospitalisation records (Hospital Episode Statistics) and to area-level deprivation data (the 2019 English Index of Multiple Deprivation).

### Population

The target population was patients newly prescribed aripiprazole, olanzapine, quetiapine, or risperidone in primary care (the four most frequently prescribed antipsychotics in the UK) [4]. On the date of first prescription for the SGA (index date), patients were included if they: were aged 18 to 99 years; had an SMI diagnosis recorded in primary care; were registered at the primary care practice for at least six months; and had results of at least one blood test for lipids or glycated haemoglobin (HbA1c) recorded in the last two years. We excluded patients who had been first prescribed the SGA prior to the study period; prescribed >1 antipsychotic on the index date (or a long-acting injectable antipsychotic in the prior 90 days) and who had a dementia diagnosis recorded in primary care. SMI diagnosis was defined as a recorded code indicating diagnosis of schizophrenia, bipolar disorder, or other non-organic psychoses (e.g., schizoaffective disorders, delusional disorder, psychotic episodes, non-organic psychosis not otherwise specified).

Patients meeting eligibility criteria entered the study on their index date between 1 January 2005 to 31 December 2017. Patients completed the study at the earliest of completion of two-year follow-up (final follow-up, 31 December 2019); end of primary care registration; death; or the last data collection date from the primary care practice.

### Data management

To identify prescriptions of antipsychotics[4] and metformin (Supplementary Table 1), we developed a search strategy (which considered generic and brand names) to identify relevant product codes in CPRD product dictionaries. We used the resulting code lists to extract data on the relevant medications from patient prescription records.

To characterise the cohort, we extracted the following demographic and baseline characteristics: sex, ethnicity, SMI diagnosis, age (at first SMI diagnosis, first antipsychotic prescription, and at cohort entry), geographical region, area-level deprivation, comorbidities (cerebrovascular disease, dyslipidaemia, myocardial infarction, hypertension, liver disease, renal disease, diabetes, alcohol misuse, substance misuse, polycystic ovarian syndrome), concomitant medication prescriptions (antidepressants, lipid-regulating medications, insulin), biochemical parameters (HbA1c, random glucose), smoking status, body mass index (BMI) and body weight.

For patients in England, if ethnicity was not coded in the primary care record, we supplemented it with ethnicity data from linked HES records, where available. If a patient had more than one ethnicity category recorded, we used the most frequent (or most recent, if frequencies were equal). Geographic region refers to the patient’s registered primary care practice and includes Northern Ireland, Scotland, Wales, and nine regions across England as per the Office for National Statistics categories. For patients in England, relative deprivation was derived from linked small area-level data using the 2019 English Index of Multiple Deprivation, based on either the patient’s residential postcode or, if unavailable, the primary care practice postcode. If a patient had multiple SMI diagnoses were recorded, the most recent diagnosis category as at the index date was used, retaining the first diagnosis date. Binary indicators were used to define antidepressant (as categorised by the British National Formulary) and insulin prescriptions in the two years prior to the index date. BMI (kg/m^2^) was categorised as underweight (<18.5), healthy weight (≥18.5 to <25), overweight (≥25 to <30), and obese (≥30).

### Missing data

Missing weight values and covariate data were handled using multiple imputation by chained equations using the ‘mice’ package in R. The imputation model included baseline, outcome, and auxiliary variables and we assumed data were missing at random conditioned on the observed variables in the model. We generated 25 imputed datasets and pooled results across imputed datasets according to Rubin’s rules.

### Statistical analysis

All analyses were conducted in R (Version 4.4.1). The full analytical code is available at: https://github.com/Alvin-RB/antipsychotic_metformin_coprescription.

To describe the incidence and prevalence of metformin prescription (objective 1), we first calculated the number of patients newly prescribed metformin when initiating the SGA in the following time-windows: ≥ 1 month before the incident SGA, ≤ 1 month before to ≤2 years after incident SGA, ≥ 2 years after the incident SGA. We then estimated cumulative incidence at the patient level and period prevalences at the time-period level. For cumulative incidence, we used the Kaplan-Meier method to model the time to first metformin prescription among patients not previously prescribed metformin more than one month prior to the index date. To ensure that patients who were prescribed metformin on, or within one month prior to, the index date were included, we set their follow-up time at 0.5 days. For all other patients, follow-up time was censored at the earliest of: first metformin prescription, date of death, end of primary care registration, last data collection data from the primary care practice, or completion of two-years follow-up from index date. To examine prevalence over time, we calculated annual period prevalences (standardised per 1,000 patients); the numerator was the total number of unique patients with at least one metformin prescription each year 2005-2017 over a denominator of all eligible patients alive and remaining in follow-up in the given year.

To compare the characteristics of patients who were prescribed metformin and those who were not (objective 2), we stratified the key demographic and baseline characteristics of the two groups by metformin exposure status. The first group consisted of patients who had never been prescribed metformin, referred to as the SGA-only group, whereas the second group included patients who had been prescribed metformin ≤1 month prior to up to ≤2 years after the initiation of the SGA, referred to as the SGA+metformin group. Similarly, to investigate the potential rationale for prescribing, we compared the characteristics of patients in the SGA+metformin group with a recorded potential indication for metformin (i.e., diabetes and/or PCOS) versus those without a recorded indication. Descriptive statistics were used to characterise the cohorts.

To describe changes in weight over time among patients prescribed and not prescribed metformin (objective 3), we plotted descriptive line graphs of the mean weight at baseline and at six-month, one- year and two-year follow-up, as well as the percentage change from baseline at each follow-up time- point (all with 95% confidence intervals [CIs]). Amongst patients in the SGA+metformin group, we included only those prescribed metformin ≤1 month prior to up to ≤3 months after the index date (i.e., those first prescribed metformin in relatively close temporality to SGA initiation). This time range was selected as the first outcome time-point was six months and we hypothesised that metformin initiated closer to the outcome, i.e. >3 months, would not allow sufficient time for the medication to exert an effect. In these primary analyses, we replaced missing values with multiple imputation (see ‘Missing data’) but we also explored the impact of missing data through an observed values analysis.

To estimate the potential effect of metformin on weight change, we constructed linear regression models to estimate the mean difference in weight (kg) at six months, one year, and two years post- index date. The models included an indicator for metformin status as the main exposure and weight (kg) as the outcome. In an initial model (Model 1), we adjusted for the baseline weight. In Model 2, we adjusted for all the confounders identified using a directed acyclic graph [29](Supplementary Figure 2). The directed acyclic graph was created to determine confounders of the relationship between metformin prescription and weight change. The variables included in the models were baseline weight (kg), antipsychotic medication, sex, age at index date, ethnicity, social deprivation, prior diagnosis of diabetes, and prior diagnosis of PCOS. Models were stratified by sex, as PCOS - a key confounder - only affects females. The models were parameterised with linear terms for all variables and additional quadratic terms for continuous variables. As above - amongst patients in the SGA+metformin group, we included only those prescribed metformin ≤1 month prior to up to ≤3 months after the index date.

## RESULTS

### Objective one: Prevalence and incidence of metformin co-prescription

Among 26,537 patients diagnosed with SMI newly initiating aripiprazole, olanzapine, quetiapine, or risperidone, 4652 were ever prescribed metformin and 21,885 were not. A total of 696 were first prescribed metformin between ≤1 month before to ≤2 years after first prescribing of the SGA, 2873 were prescribed metformin ≥ 1month before the SGA, and 1083 were first were prescribed metformin ≥2 years after the incident SGA (Figure 1, Supplementary Figure 1). Among the patients not prescribed metformin within one month prior to the date of first prescription of the SGA (n=23,664), the cumulative incidence of first metformin prescription at one year was 1.9% (95% CI, 1.8 to 2.1) and, at two years, 3.3% (95% CI, 3.0 to 3.5) (Supplementary Figure 3). At the time-period level, period prevalence increased from 13.1 per 1,000 patients (95% CI, 6.5 to 19.8) in 2005 to 58.4 (95% CI, 52.2 to 64.5) in 2017 (Supplementary Figure 4).

**FIGURE 1:**
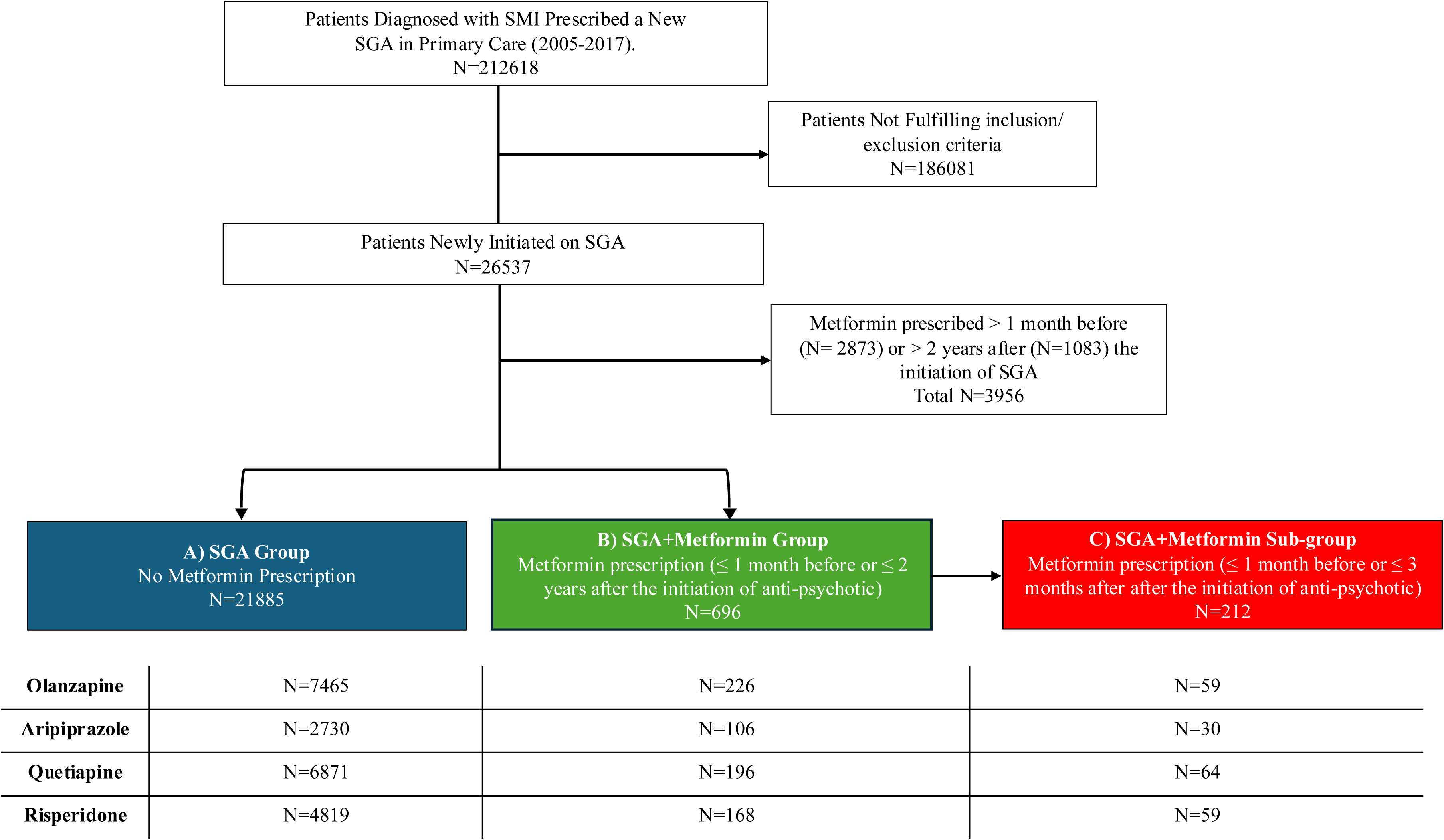
Study flow diagram.

### Objective two: Characteristics of patients co-prescribed and not co-prescribed metformin

Characteristics of the SGA-only compared to the SGA+metformin group are shown in Table 1. The distribution of the types of SGAs prescribed was similar; olanzapine was the most common. The SGA-only group had a higher proportion of white individuals (84.24%) compared to the SGA+metformin group (75.23%) while the SGA+metformin group has higher percentages of Asian (11.99% vs. 6.42%) and Black (10.12% vs. 6.23%) individuals. The SGA+metformin group had higher levels of social deprivation, with 34.41% residing in the most deprived IMD quintile compared to 27.37% in the SGA-only group. With respect to SMI diagnosis, schizophrenia (25.29% vs 18.87%) was more common in the SGA+metformin group, while other non-organic psychoses (38.25% vs 33.91%) and bipolar disorder (42.88% vs 40.80%) was more common in the SGA-only group. The groups had similar ages at first SMI diagnosis, first antipsychotic prescription, and at cohort entry and there were limited geographical differences.

**TABLE 1:** Characteristics of patients prescribed a second-generation antipsychotic (only) compared to patients prescribed a second-generation antipsychotic and metformin (**≤**1 month before to **≤**2 years after SGA prescription).

| Characteristic | SGA Only<br>N = 21,885 | SGA+Metformin<br>N = 696 |
| --- | --- | --- |
| **Gender, N (%)** |  |  |
| Female | 12,130 (55.43%) | 372 (53.45%) |
| Male | 9,755 (44.57%) | 324 (46.55%) |
| **Age, Mean (SD)** |  |  |
| at index date | 53.73 (18.00) | 53.57 (14.28) |
| at first SGA prescription | 50.95 (17.87) | 50.51 (14.34) |
| at first SMI diagnosis | 45.64 (19.01) | 44.90 (15.75) |
| **Ethnicity, N (%)** |  |  |
| Asian | 1,256 (6.42%) | 77 (11.99%) |
| Black | 1,218 (6.23%) | 65 (10.12%) |
| Mixed/Other | 608 (3.11%) | 17 (2.64%) |
| White | 16,476 (84.24%) | 483 (75.23%) |
| Unknown | 2,327 | 54 |
| **SGA initiated at index date, N (%)** |  |  |
| Aripiprazole | 2730 (12.47%) | 106 (15.23%) |
| Olanzapine | 7465 (34.11%) | 226 (32.47%) |
| Risperidone | 4819 (22.02%) | 168 (24.14%) |
| Quetiapine | 6871 (31.40%) | 196 (28.16%) |
| **SMI diagnosis, N (%)** |  |  |
| Bipolar disorder | 9,384 (42.88%) | 284 (40.80%) |
| Other non-organic psychoses | 8,372 (38.25%) | 236 (33.91%) |
| Schizophrenia | 4,129 (18.87%) | 176 (25.29%) |
| **Geographical region, N (%)** |  |  |
| East Midlands | 437 (2.00%) | 13 (1.87%) |
| East Of England | 911 (4.16%) | 26 (3.74%) |
| London | 4,217 (19.27%) | 162 (23.28%) |
| North East | 676 (3.09%) | 18 (2.59%) |
| North West | 3,597 (16.44%) | 122 (17.53%) |
| Northern Ireland | 517 (2.36%) | 10 (1.44%) |
| Scotland | 1,608 (7.35%) | 40 (5.75%) |
| South East | 3,416 (15.61%) | 94 (13.51%) |
| South West | 1,903 (8.70%) | 59 (8.48%) |
| Wales | 1,220 (5.57%) | 44 (6.32%) |
| West Midlands | 2,780 (12.70%) | 88 (12.64%) |
| Yorkshire & The Humber | 603 (2.76%) | 20 (5.63%) |
| **2019 English IMD quintile, N (%)** |  |  |
| 1 (Least Deprived) | 2,568 (14.23%) | 57 (9.66%) |
| 5 (Most Deprived) | 4,939 (27.37%) | 203 (34.41%) |
| Unknown | 3,842 | 106 |
| **Baseline Weight (kg), Mean (SD)** | 76.83 (19.22) | 90.39 (23.47) |
| **Baseline BMI<sup>2</sup>, N (%)** |  |  |
| Underweight | 615 (3.69%) | 7 (1.22%) |
| Healthy | 5,849 (35.10%) | 79 (13.72%) |
| Overweight | 5,539 (33.24%) | 146 (25.35%) |
| Obese | 4,662 (27.97%) | 344 (59.72%) |
| *Unknown* | 5,220 | 120 |
| **Comorbidities, N (%)** |  |  |
| Cerebrovascular disease | 1,439 (6.58%) | 43 (6.18%) |
| Myocardial infarction | 710 (3.24%) | 27 (3.88%) |
| Liver disease | 471 (2.15%) | 17 (2.44%) |
| Renal disease | 2,235 (10.21%) | 48 (6.90%) |
| Hypertension | 5,951 (27.19%) | 273 (39.22%) |
| Dyslipidaemia | 3,983 (18.20%) | 169 (24.28%) |
| Diabetes | 2,072 (9.47%) | 339 (48.71%) |
| Polycystic Ovarian Syndrome | 286 (2.36%) | 21 (3.02%) |
| **Other medications prescribed in prior 2y, N (%)** |  |  |
| Antidepressants | 12,972 (59.27%) | 427 (61.35%) |
| Lipid-regulating medications | 5,360 (24.49%) | 293 (42.10%) |
| Insulin | 287 (1.31%) | 25 (3.59%) |
| **Prior Exposure to Substances, N (%)** |  |  |
| Alcohol misuse | 2,436 (11.13%) | 65 (9.34%) |
| Substance misuse | 1,823 (8.33%) | 51 (7.33%) |
| Ex-Smoker | 3,700 (16.99%) | 104 (15.03%) |
| Current Smoker | 9,383 (43.07%) | 314 (45.38%) |
| **Biochemical parameters,<sup>3</sup> Mean (SD)** |  |  |
| HbA1c | 39.06 (10.35) | 54.65 (20.02) |
| *Unknown* | 14,968 | 264 |
| Glucose | 5.30 (1.58) | 7.61 (3.70) |
| *Unknown* | 4,666 | 112 |

SGA, second generation antipsychotic; IMD, index of multiple deprivation; BMI, body mass index; HbA1c, glycated hemoglobin.
Percentages are of patients with non-missing data.
<sup>1</sup> Percentages out of total number of females.
<sup>2</sup> BMI (kg/m<sup>2</sup>) categories: underweight (<18.5), healthy weight (≥18.5 to <25), overweight (≥25 to <30), obese (≥30).
<sup>3</sup> Biochemical parameters are defined according to test result values recorded on or within the two years prior to the index date, using the value closest to the index date. For values requiring a blood test for measurement, values recorded up to seven days after the index date were also considered on the assumption that results might relate to the date on which test results were received, rather than the date on which the blood test was taken.

The SGA+metformin group had a higher mean (SD) baseline weight (90.39 [23.47] kg) compared to the SGA-only group (76.83 [19.22] kg). With regards to BMI, 27.97% of the SGA-only patients were obese. In the SGA+metformin group, 59.72% of patients were considered obese. The SGA+metformin group had more patients with dyslipidaemia (24.28% vs. 18.20%), hypertension (39.22% vs. 27.19%) and diabetes (48.71% vs. 9.47%). Across the groups, the number with previous myocardial infarction, cerebrovascular disease, renal disease, and liver disease were broadly similar, as was smoking status, alcohol misuse and substance misuse. A higher proportion in the SGA+metformin group were prescribed lipid-lowering drugs (42.10% vs. 24.49%) and insulin (3.59% vs. 1.31%), but the groups had similar proportions of patients prescribed antidepressants.

When stratified by recorded potential metformin indication, 355 (51.0%) of the SGA+metformin group had a record of diabetes and/or PCOS and 341 (49.0%) did not. The 341 patients who did not have a recorded diagnosis of diabetes or PCOS were more likely to be overweight (65.50% vs 55.03%) than those with a recorded potential metformin indication (Supplementary Table 2).

Objective three: Changes in body weight over time.

Of the 696 patients in the SGA+metformin group, 212 were prescribed metformin ≤1 month prior to up to ≤3 months after the index date (i.e., first prescribed metformin in relatively close temporality to SGA initiation). These 212 patients were included in the analyses for this objective and compared to the 21,885 patients who were prescribed SGA-only.

By two years post-SGA initiation, the mean body weight in the SGA only group had changed by +4.16% (95% CI, -1.26 to +9.58) compared to -0.65% (95% CI, -4.26 to +2.96) in the SGA+metformin group (Figure 2, Supplementary Table 3, and Supplementary Figure 5). Patients in the SGA+metformin group persisted to have greater absolute body weight than those in the SGA only group over follow-up; change over time in the absolute values are shown in Figure 2, Supplementary Table 3, and Supplementary Figure 5.

**FIGURE 2:**
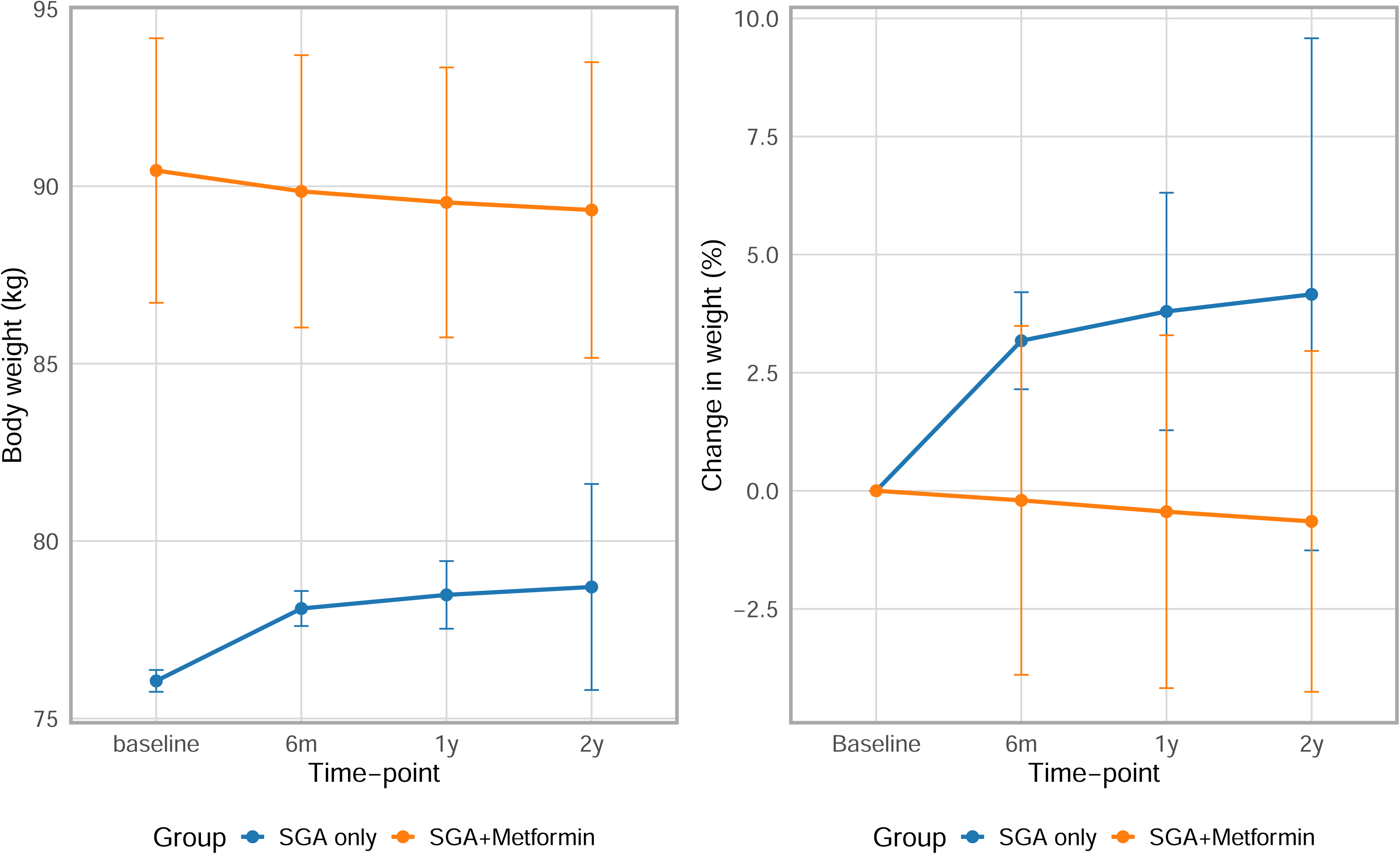
Mean absolute weight and Mean percentage change in weight over time in patients prescribed SGA only (N= 21,885) versus those prescribed SGA+Metformin (initiated ≤ 1 month before or ≤ 3 months after SGA prescription) (N=221). SGA, second-generation antipsychotic. Missing data were replaced using multiple imputation.

Stratified by SGA, patients prescribed all four SGAs (without metformin) experienced weight gain over time. In contrast, in the SGA+metformin group - weight remained similar or decreased across the four SGAs (Supplementary Figures 6-9).

When adjusting for baseline weight, the mean difference in body weight at two-years with metformin co-prescription was -2.04 kg (95% CI, -4.55 to -0.47) among females and -3.02 kg (95% CI, -5.84 to - 0.20) among males (Figure 3). After adjusting for all specified confounders, these differences attenuated slightly; the mean difference in body weight at two years with co-prescription was -1.48 kg (95% CI, -4.03 to 1.07) among females and -1.84 kg (95% CI, -4.67 to 0.98) among males (Figure 3).

**FIGURE 3:**
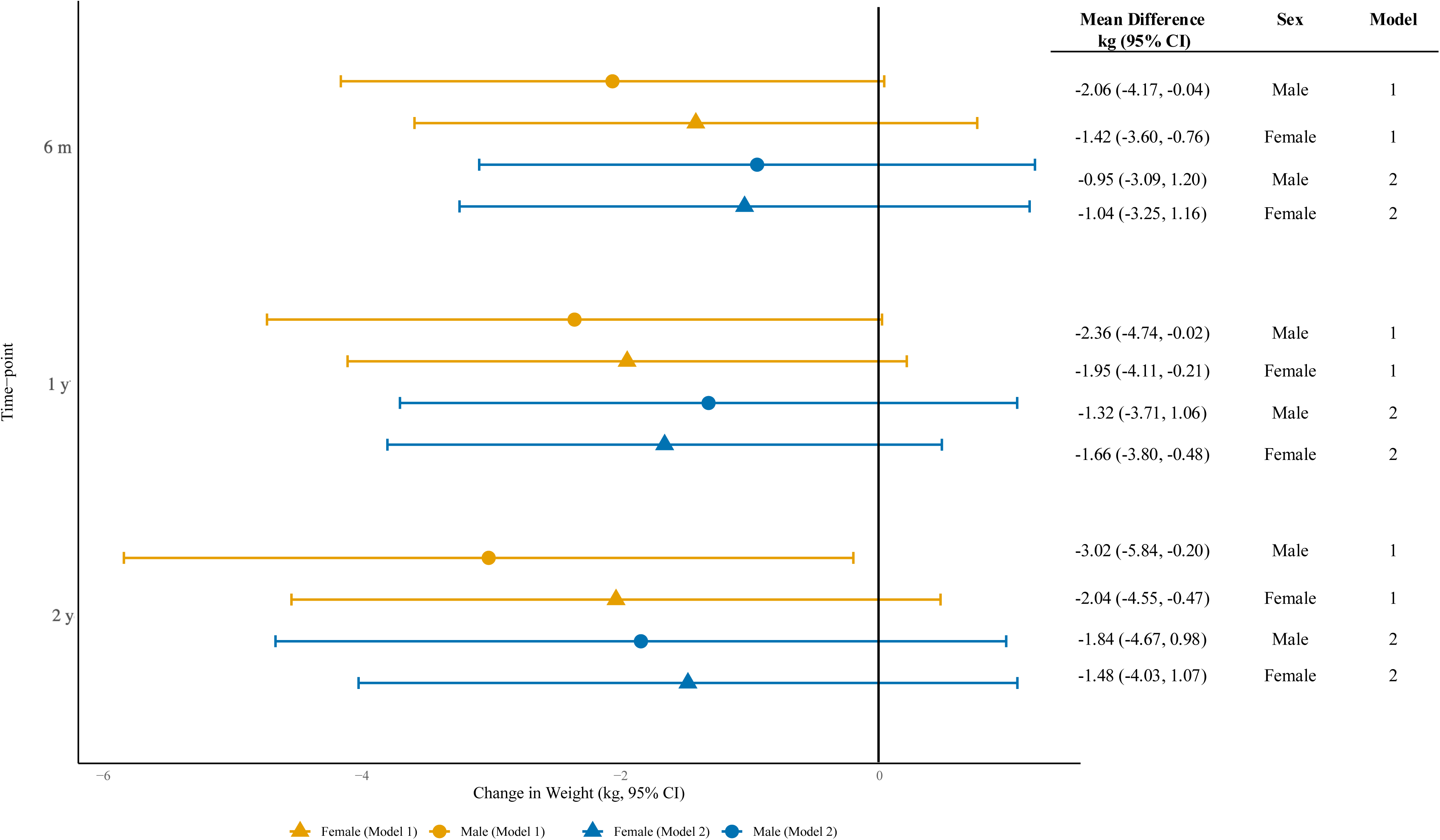
Forest plot of the estimated effect of metformin on change in weight at six months, one year and two years post-index date. Model 1 is adjusted for baseline weight (linear and quadratic terms). Model 2 is adjusted for the following covariates: baseline weight (linear and quadratic terms), second-generation antipsychotic type, age at baseline (linear and quadratic terms), ethnicity, socioeconomic deprivation index, prior diabetes, PCOS (for females only). Separate models were fitted for each sex. For each model, weight change was regressed on a metformin indicator variable.

## DISCUSSION

In a large cohort of patients diagnosed with SMI, we found that few patients were co-prescribed metformin alongside the four most frequently prescribed antipsychotics in the UK (3.3% two-year cumulative incidence). A substantial proportion of patients who were prescribed metformin had other recorded potential indications (i.e., diabetes or PCOS), suggesting that metformin is not being prescribed for the prevention or management of AIWG. This means that the group of patients receiving metformin specifically for AIWG is likely to be even smaller - we approximate this to be around 1% of the study cohort, raising concerns about unmet need for metabolic risk management in this population. Perhaps promisingly, we observed that the period prevalence of metformin co-prescription over time (from 13.1 to 58.4 per 1,000 patients), however, the absolute numbers remained very low. There did not appear to be any impact on period prevalences associated with the BAP guidance published in 2016 [22]. Our data also highlights that patients who were co-prescribed metformin were more likely to be from deprived areas and have obesity and comorbid conditions, such as diabetes and hypertension.

Through descriptive analyses, we observed a lack of weight gain in patients who were prescribed metformin ≤1 month before to up to ≤3 months after SGA initiation compared to those on SGA therapy alone. After adjustment for multiple confounders, which we identified using a direct acyclic graph, the mean difference in body weight at two years with co-prescription was -1.48 kg (95% CI, -4.03 to 1.07) among females and -1.84 kg (95% CI, -4.67 to 0.98) among males; however, confidence intervals were wide - likely driven by the relatively small number of patients co-prescribed metformin. This means that we could not draw firm conclusions regarding the effect of metformin on weight change; further well-powered studies would be needed to reliably estimate the effect of metformin co-prescription in UK practice[23]. However, it is important to note that patients prescribed metformin had a higher baseline weight and were more likely to have an overweight or obese BMI, potentially leading to increased non-pharmacological interventions, such as dietary and exercise advice, as well as active weight management efforts, for which we did not have data on.

Several factors could contribute to the under-utilisation of metformin. Prescribers may be deterred by potential side effects such as abdominal pain, nausea, vomiting, and vitamin B12 deficiency[15], particularly when combined with the gastrointestinal side effects associated with antipsychotic medications[30]. In addition, alcohol enhances the hypoglycaemic effect of metformin and increases the risk of lactic acidosis and therefore metformin is contraindicated for people with comorbid harmful use of alcohol. It is recommended to monitor renal function annually and vitamin B12 levels in those who are at a higher risk of developing a deficiency[15], patients who are considered less likely to engage in monitoring may be less likely to be prescribed metformin. Furthermore, AIWG is not explicitly listed as an indication in the British National Formulary, creating another barrier to its prescription for this purpose[15]. Furthermore, the NICE surveillance review states that clinicians should “consider” metformin as an adjunctive therapy[24]. We were not able to ascertain how many patients had metformin considered and then subsequently not prescribed. In addition, the guidelines are unclear as to whether metformin should be used as a prophylactic or management strategy for AIWG. Therefore, more clarity is needed to appropriately guide patients and clinicians. Finally, shared care of patients diagnosed with SMI between primary care and psychiatric services can lead to assumptions that the other party is managing the cardiometabolic risks of antipsychotic medications, resulting in missed interventions; a collaborative approach between both services is crucial to proactively monitor and address metabolic risks. There is also ongoing interest into other potential candidates for the management of AIWG, in particular, GLP-1 inhibitors (liraglutide and exenatide) have been highlighted by NICE[24]. Further research is needed to evaluate the real-world prescribing patterns and efficacy of GLP-1 inhibitors in patients with SMI.

### Strengths and limitations

This study has several strengths. To our knowledge, this is the first study to analyse metformin co-prescription in individuals diagnosed with SMI initiating SGA therapy in UK primary care. Data generated from our study can be used to inform subsequent guideline implementation with the potential to address cardiometabolic risks in an under-served population. Another key strength lies in the large cohort of patients diagnosed with SMI from a data source representative of the UK population[28]. Moreover, the 12-year study period provided an opportunity to identify longitudinal prescribing trends, rather than limiting the findings to a single temporal snapshot.

Nonetheless, limitations must be acknowledged. First, the data set included prescriptions issued in primary care only, thereby not including metformin prescribed in hospital-based outpatient clinics. Despite hospital-based outpatient clinics often initiating prescriptions, primary care will be responsible for ongoing repeat prescriptions and so ultimately these would be represented in our data. Second, we did not have information on individual patient adherence, however, if patients are receiving repeat prescriptions, one might infer adherence. Third, the study period concluded in 2019, and thus did not encompass the COVID-19 pandemic, which may have impacted prescribing patterns. Fourth, although approximately 700 patients commenced metformin after SGA initiation, restricting the analysis to those who initiated metformin closer to SGA initiation reduced the sample size to 212, limiting statistical power and preventing firm conclusions about metformin’s effect on weight change to be drawn; however, this still compared favourably to the largest trial of metformin in this population (N=116). Finally, our study focused on UK prescribing practices and may not generalise to other countries. Further studies are needed to explore the role of metformin co-prescription in other healthcare systems.

## Conclusion

Overall, these findings highlight that metformin is highly underutilised, despite its efficacy and clinical guidelines. Additional research is needed to investigate the patient groups that would derive the most benefit from this intervention. Additionally, there is a need for clear guidance on the implementation of guidelines to achieve evidenced-based care as well as to define the roles and responsibilities of clinicians in managing patients who experience the cardiometabolic sequelae of SGAs.

### Clinical Implications

Our findings suggest a need for a proactive approach to cardiometabolic risk management through improved collaboration between primary care and psychiatric services. Enhancing prescriber awareness of metformin’s potential benefits, alongside addressing barriers such as off-label prescribing challenges and patient adherence, is warranted. Future research should focus on the long-term efficacy of metformin for AIWG and its implementation in routine clinical practice.

## Data Availability

Data was accessed via Clinical Practice Research Datalink (CPRD) under approved protocol no. 21_000729. Authors cannot share the data directly. However, data can be accessed directly from Clinical Practice Research Datalink (CPRD) following approval and licensing

https://github.com/Alvin-RB/antipsychotic_metformin_coprescription

## ACKNOWLEDGEMENTS

Thank you to the patients and primary care practices contributing to the Clinical Practice Research Datalink database. This study is partly based on data from the Clinical Practice Research Datalink obtained under licence from the UK Medicines and Healthcare products Regulatory Agency. The data is provided by patients and collected by the NHS as part of their care and support. The interpretation and conclusions contained in this study are those of the author/s alone.

## Contributions

LFT, ARB, JFH, and DPJO formulated the research questions and designed the study. LFT and ARB analysed the data. LFT wrote the first draft of the manuscript and ARB, JFH, NL, DPJO, NMD critically reviewed the manuscript for important intellectual content. ARB, NL, LFT, JFH and DPJO had full access to the data. All authors approved the final version to be published and agree to be accountable for all aspects of the work in ensuring that questions related to the accuracy or integrity of any part of the work are appropriately investigated and resolved.

## Competing interest statement

JFH has received consultancy fees from Wellcome Trust and funding grants from juli Health. All other authors declare no potential competing interests.

## Funding statement

ARB is funded by the Wellcome Trust through a PhD Fellowship in Mental Health Science. This research was funded in whole or in part by the Wellcome Trust. For the purpose of Open Access, the author has applied a CC BY public copyright licence to any Author Accepted Manuscript (AAM) version arising from this submission.

DPJO is supported by the University College London Hospitals NIHR Biomedical Research Centre and the NIHR North Thames Applied Research Collaboration. This funder had no role in study design, data collection, data analysis, data interpretation, or writing of the report. The views expressed in this article are those of the authors and not necessarily those of the NHS, the NIHR, or the Department of Health and Social Care.

JFH is supported by UKRI grant MR/V023373/1, the University College London Hospitals NIHR Biomedical Research Centre and the NIHR ARC North Thames.

NMD is supported via a Norwegian Research Council 295989 and the UCL Division of Psychiatry.

LFT is supported by NHS England.

NL is supported by a HDR UK personal fellowship (Big Data for Complex Disease- HDR-23012). This work is affiliated to HDR UK which is funded by the Medical Research Council (UKRI), the National Institute for Health Research, the British Heart Foundation, Cancer Research UK, the Economic and Social Research Council (UKRI), the Engineering and Physical Sciences Research Council (UKRI), Health and Care Research Wales, Chief Scientist Office of the Scottish Government Health and Social Care Directorates, and Health and Social Care Research and Development Division (Public Health Agency, Northern Ireland).

## Ethical approval

The data provided by primary care practices to CPRD does not include any information that can identify patients, so individual patient consent was unnecessary. However, patients were given the option to opt out of data sharing. Ethical approval was obtained from the Independent Scientific Advisory Committee of CPRD (protocol no. 21_000729).

## Data sharing

This study’s data was accessed via Clinical Practice Research Datalink (CPRD) under approved protocol no. 21_000729. Authors cannot share the data directly. However, data can be accessed directly from Clinical Practice Research Datalink (CPRD) following approval and licensing (see https://cprd.com/ for further details).

## Notes

### Author Declarations

Ethical approval was obtained from the Independent Scientific Advisory Committee of CPRD (protocol no. 21_000729).

