## Supplementary Materials for "Co-prescription of Metformin and Antipsychotics in Severe Mental Illness: A UK Primary Care Cohort Study"

Luiza Farache Trajano, Joseph F. Hayes, Naomi Launders, Neil M. Davies,
David P. J. Osborn, Alvin Richards-Belle

### Supplementary Figures

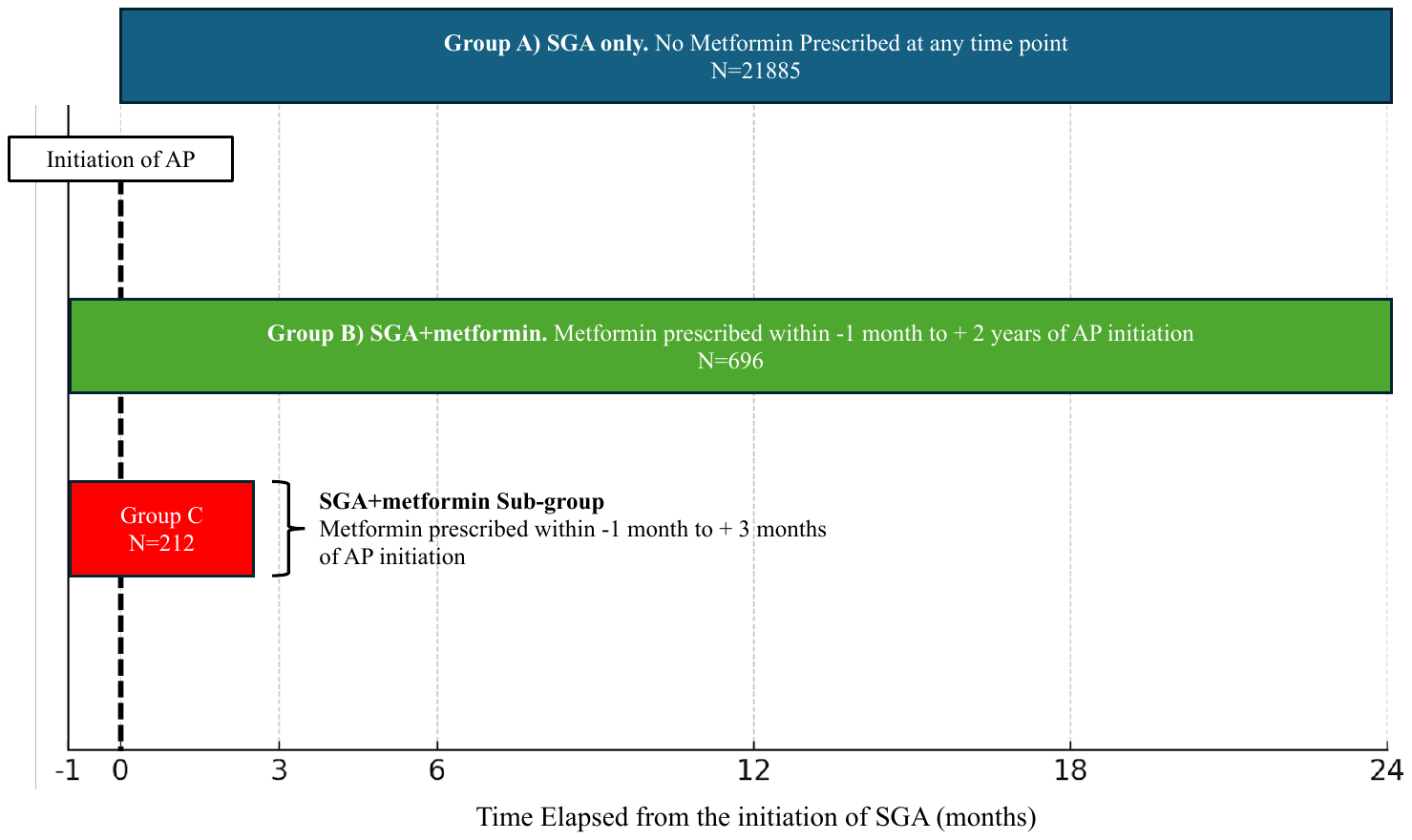

Supplementary Figure 1. Pictorial summary of the three study groups used to investigate antipsychotic and metformin co-prescription throughout this study.

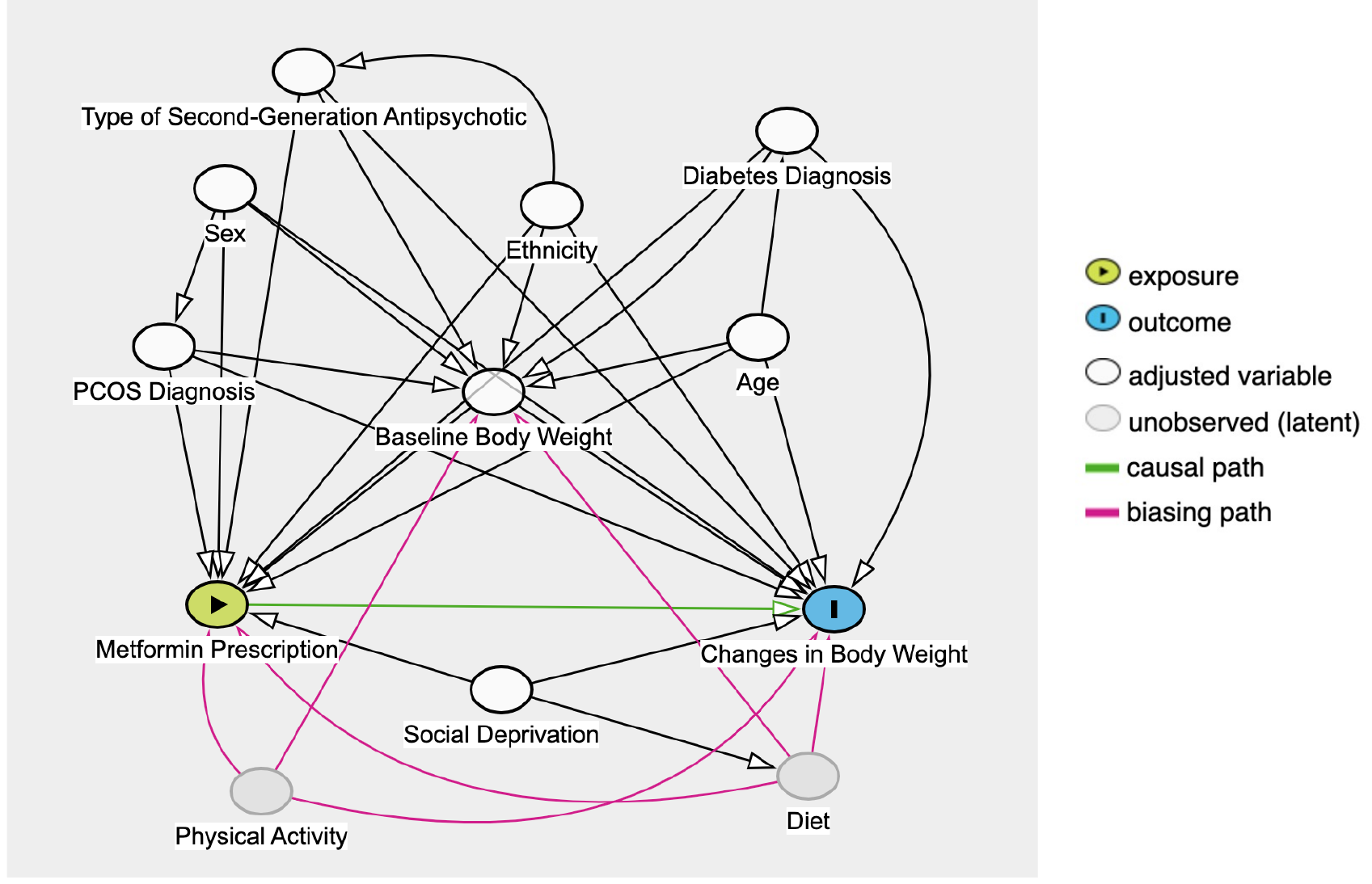

Supplementary Figure 2. Directed Acyclic Graph.

Directed Acyclic Graph (DAG) illustrating the causal pathways between metformin prescription (exposure) and change in body weight (outcome), with adjustments for multiple confounders, including baseline weight, sex, age, ethnicity, social deprivation, prior diabetes, prior PCOS, and the type of second-generation antipsychotic medication.

The green arrow represents the hypothesised causal path from metformin prescription to weight change, while black arrows depict the influence of various covariates that potentially confound the relationship between metformin prescription and weight change. Pink arrows represent biasing paths introduced by unobserved confounders such as diet and physical activity, highlighting additional sources of potential confounding in the analysis. PCOS diagnosis is a sex-specific diagnosis, affecting females only.

| **Time (years)** | **0** | **0.5** | **1** | **1.5** | **2** |
| --- | --- | --- | --- | --- | --- |
| Cumulative incidence | 0% | 1.2% (1.1%, 1.4%) | 1.9% (1.8%, 2.1%) | 2.6% (2.4%, 2.9%) | 3.3% (3.0%, 3.5%) |

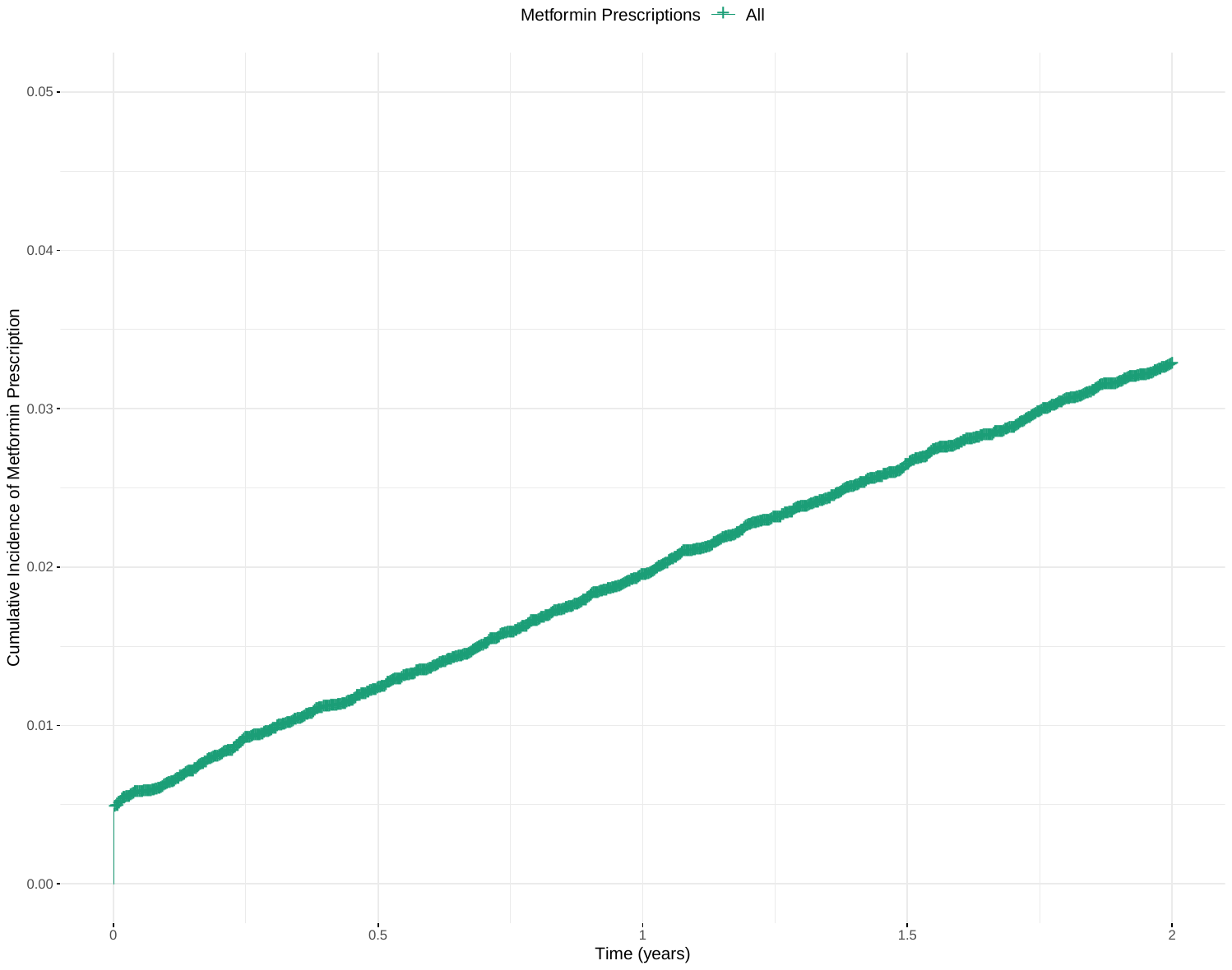

Supplementary Figure 3. Cumulative incidence of first metformin prescription.

The Kaplan-Meier method was used to model the time to first metformin prescription among patients not previously prescribed metformin more than one month prior to the index date. To ensure that patients prescribed metformin on, or within one month prior to, the index date were included, their follow-up time was set at 0.5 days. For all included patients, follow-up time was censored at the earliest of: first metformin prescription, date of death, end of primary care registration, last data collection data from the primary care practice, or completion of two-years follow-up from index date.

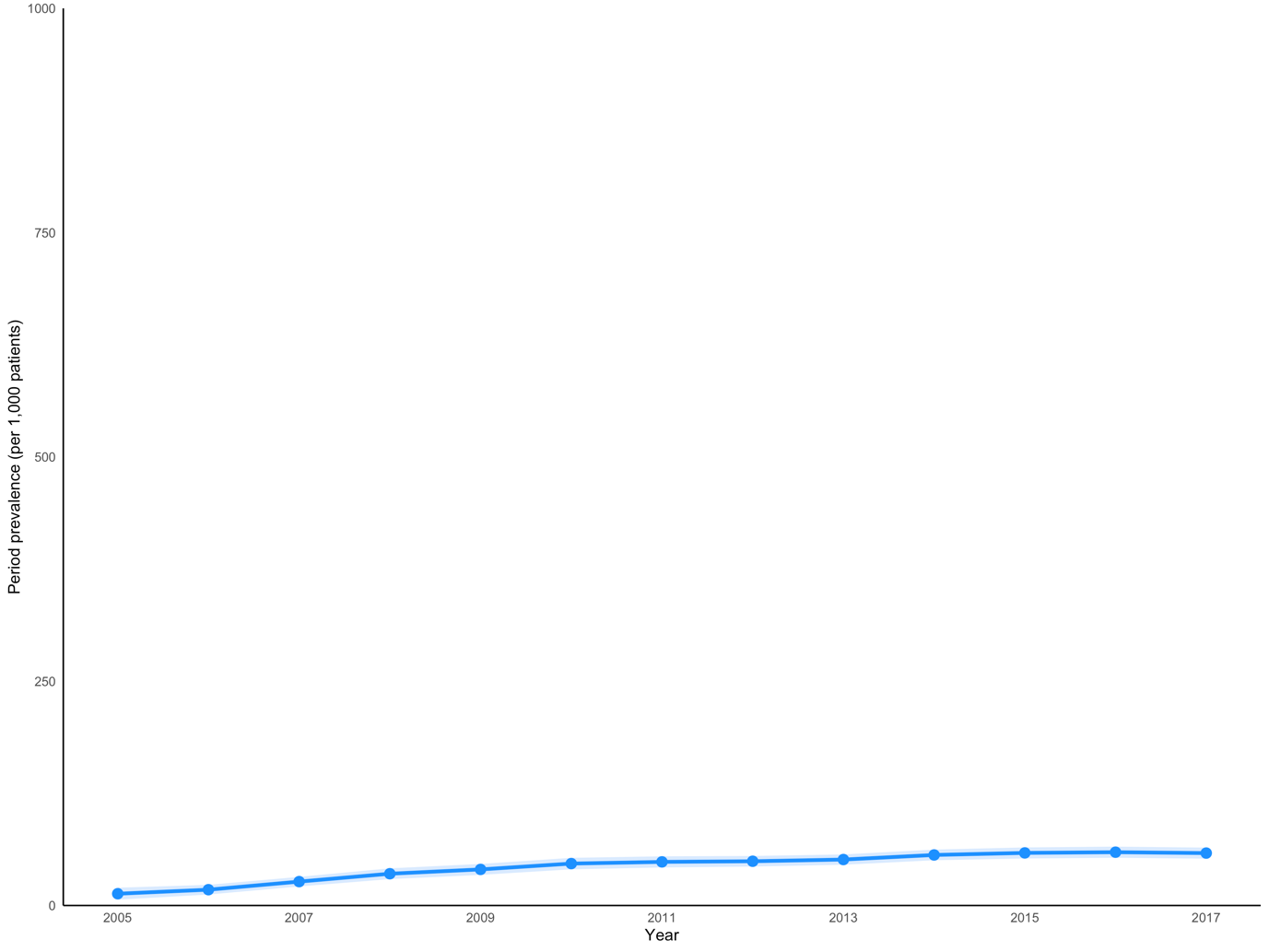

Supplementary Figure 4. Annual period prevalence of metformin prescriptions.

Rates (standardised per 1,000 patients); the numerator was the total number of unique patients with at least one metformin prescription each year 2005-2017 over a denominator of all eligible patients alive and remaining in follow-up in the given year.

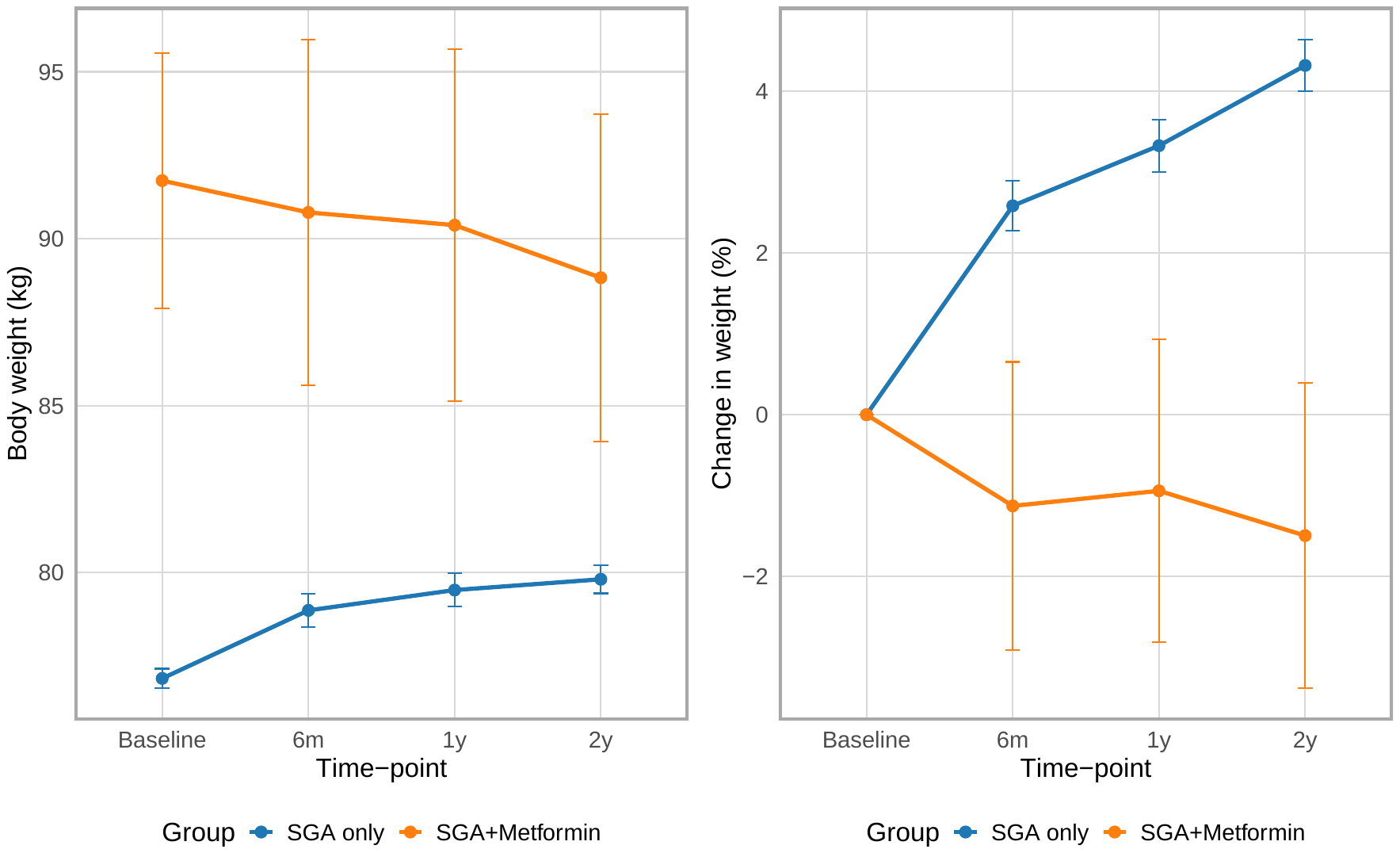

Supplementary Figure 5. Mean absolute and percentage change in weight over time in patients prescribed SGA only versus those prescribed SGA+Metformin (observed data).

This figure is based on observed data only.

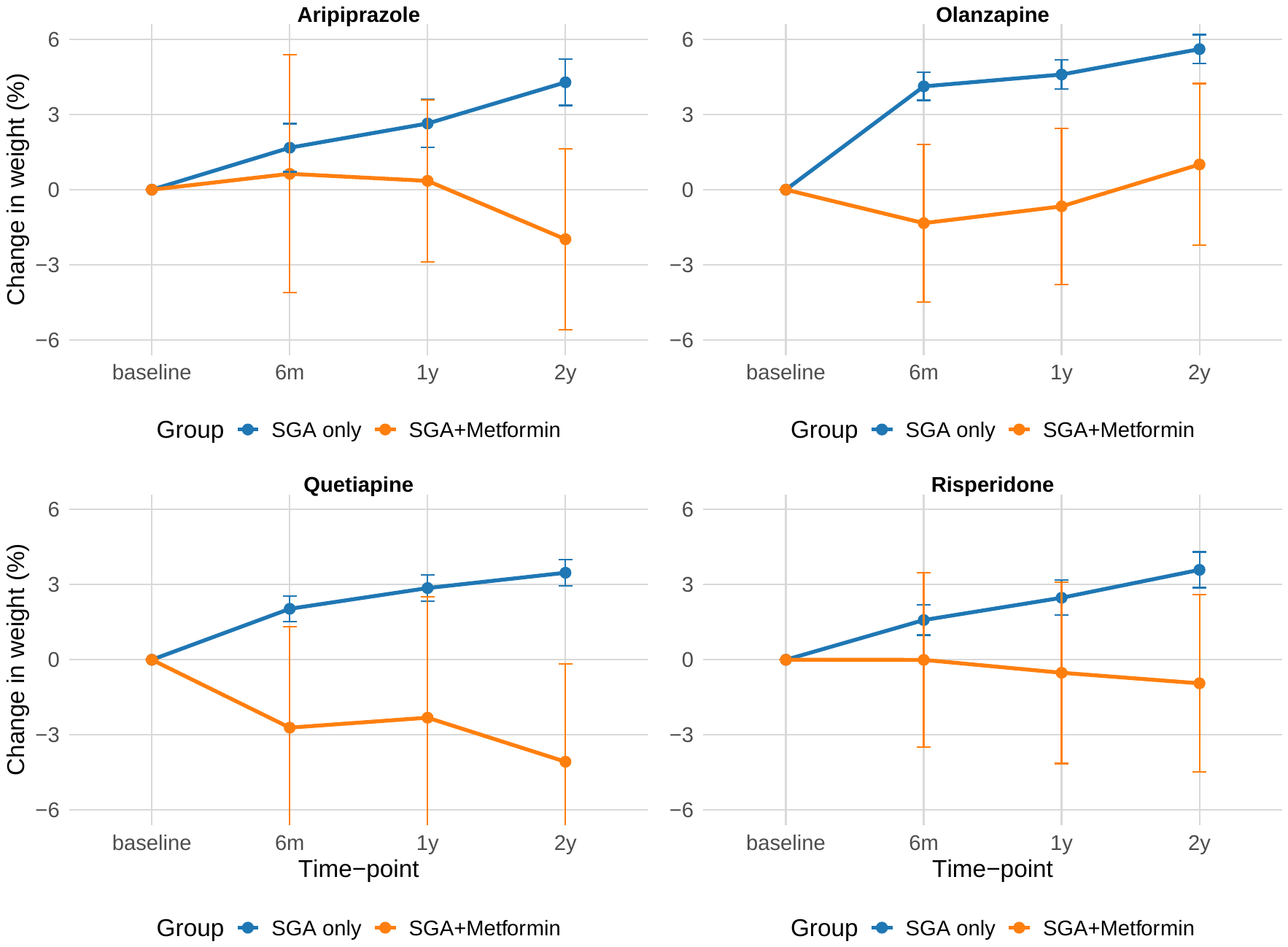

Supplementary Figure 6. Mean percentage change in weight over time in patients prescribed SGA only versus those co-prescribed SGA+Metformin, stratified by SGA (observed data).

The four panels display data for aripiprazole, olanzapine, quetiapine, and risperidone, with mean percentage weight change (%) on the y-axis and time (6 months, 1 year, 2 years) on the x-axis. This figure is based on observed data only.

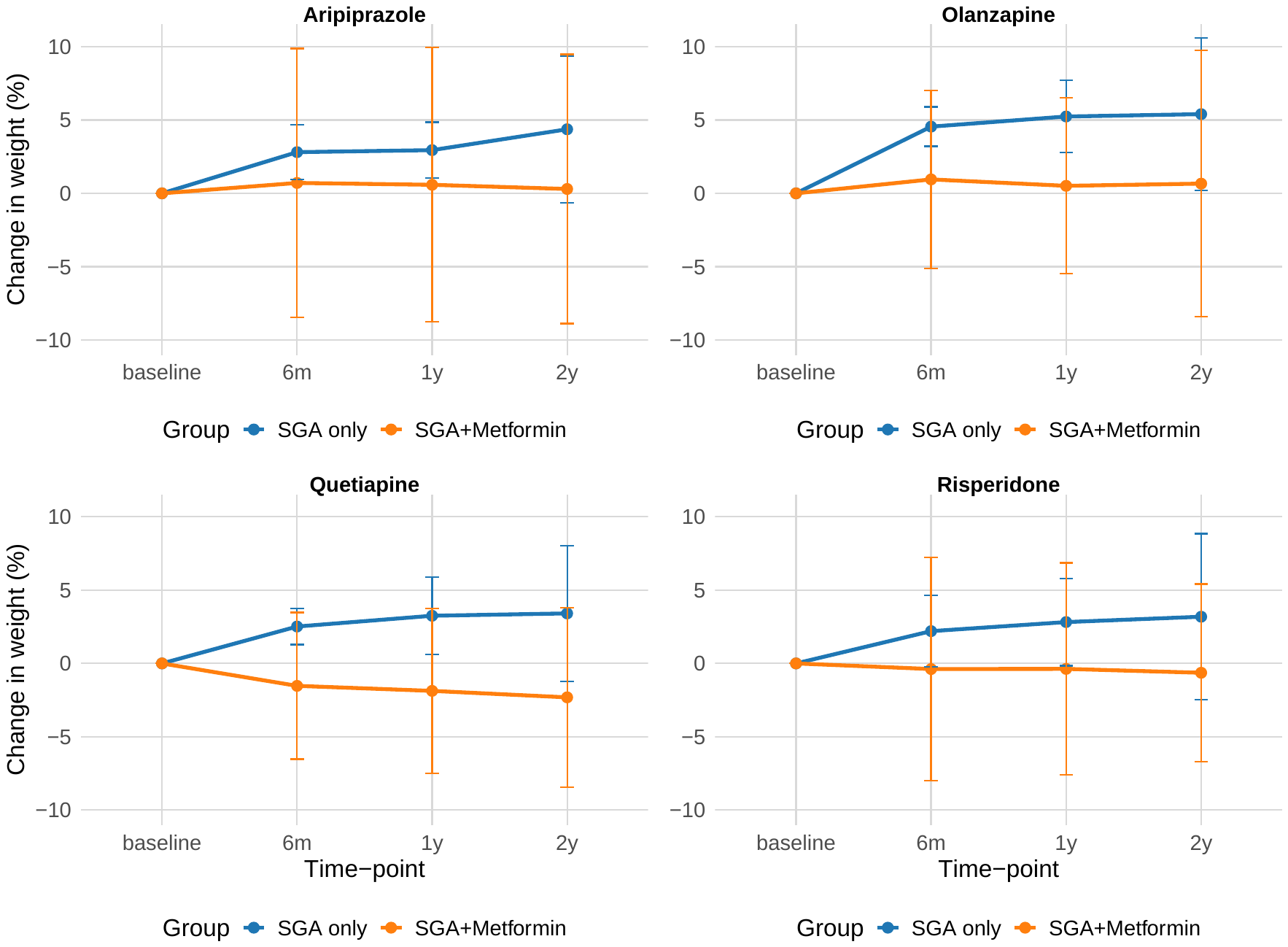

Supplementary Figure 7. Mean percentage change in weight over time in patients prescribed SGA only versus those co-prescribed SGA+ Metformin, stratified by SGA (missing data replaced using multiple imputation).

The four panels display data for Quetiapine (SGA only group N= 6871. SGA+metformin group N= 64), Olanzapine (SGA only group N= 7465. SGA+metformin group N= 226 ), Risperidone (SGA only group N= 4819. SGA+metformin group N= 168) and Aripiprazole (SGA only group N= 2730. SGA+ metformin group N= 2730), with mean body weight (kg) on the y-axis and time (6 months, 1 year, 2 years) on the x-axis. Missing values were imputed using multiple imputation.

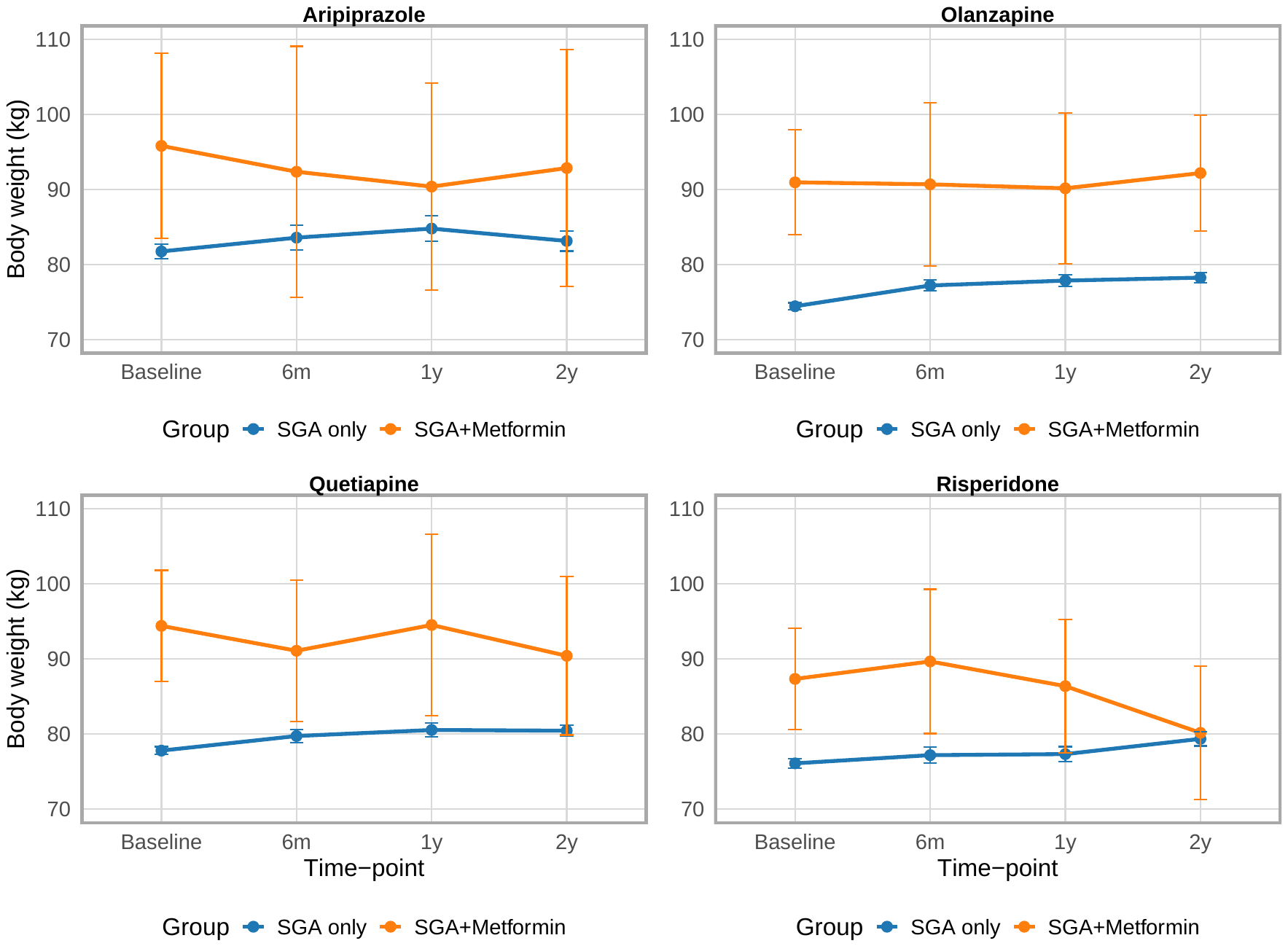

Supplementary Figure 8. Mean absolute weight (kg) over time in patients prescribed different anti-psychotics, comparing those on anti-psychotics only versus those co-prescribed anti-psychotics and Metformin (observed data).

The four panels display data for Quetiapine, Olanzapine, Risperidone, and Aripiprazole, with mean body weight (kg) on the y-axis and time (6 months, 1 year, 2 years) on the x-axis. This figure is based on observed data only.

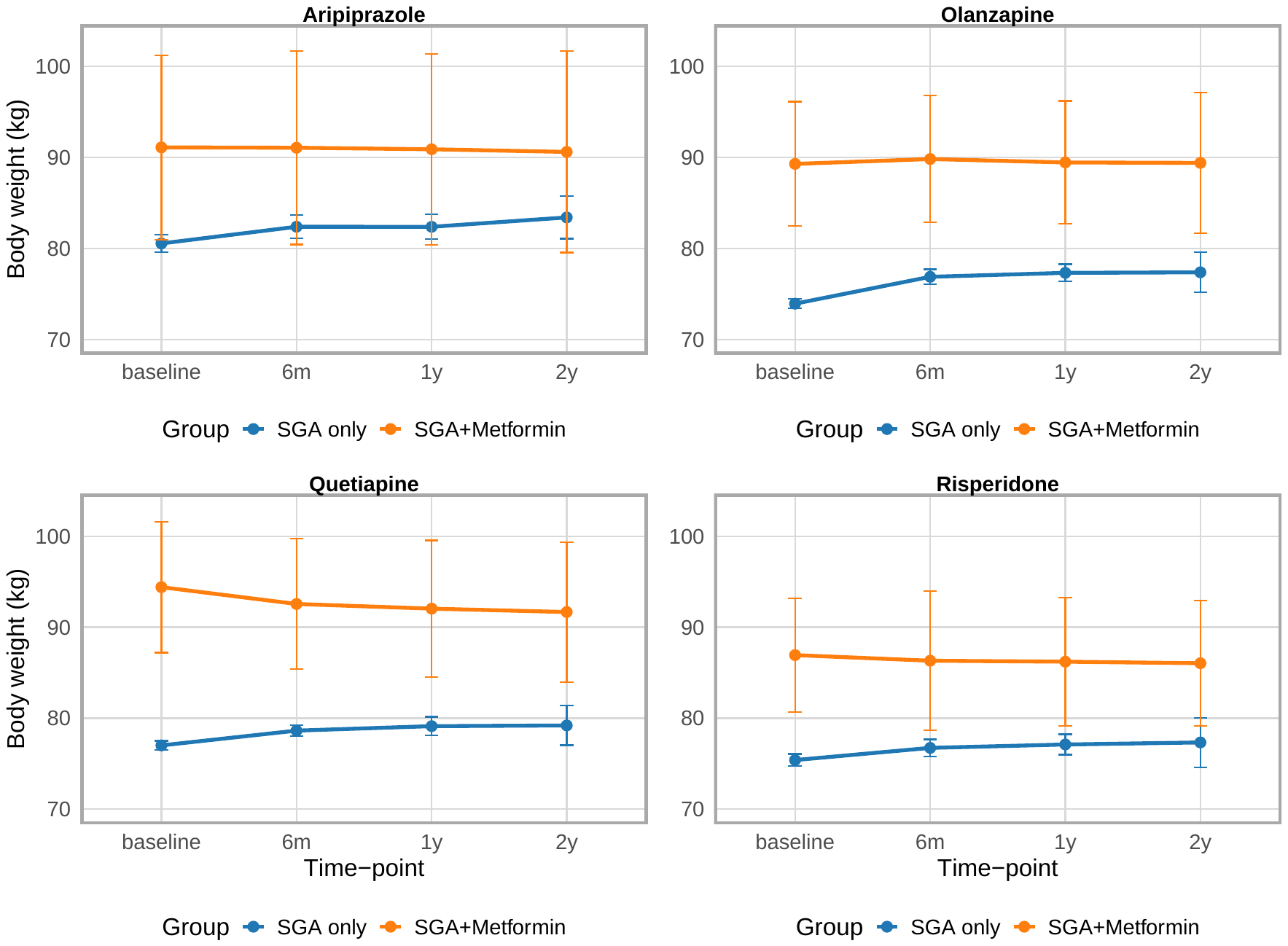

Supplementary Figure 9. Mean absolute weight (kg) over time in patients prescribed different anti-psychotics, comparing those on anti-psychotics only versus those co-prescribed anti-psychotics and Metformin (missing data replaced using multiple imputation).

The four panels display data for Quetiapine (SGA only group N= 6871. SGA+metformin group N= 64), Olanzapine (SGA only group N= 7465. SGA+metformin group N= 226), Risperidone (SGA only group N= 4819. SGA+metformin group N= 168) and Aripiprazole (SGA only group N= 2730. SGA+ metformin group N= 2730), with mean body weight (kg) on the y-axis and time (6 months, 1 year, 2 years) on the x-axis. Missing data were replaced using multiple imputation.

### Supplementary Tables

Supplemental Table 1. CPRD product code lists for antidiabetic agents containing metformin.

| **CPRD Aurum** |  |
| --- | --- |
| **Product Code** | **Product Name** |
| 644941000033117  645041000033117  896941000033112  897041000033113  2620241000033119  2995141000033110  2995241000033115  3191341000033115  3191441000033114  3200541000033116  3200641000033115  3200741000033112  3200841000033119  3228141000033117  3228241000033112  3890941000033116  3982341000033110  3982441000033116  3983941000033115  3984241000033114  4452341000033119  4452441000033113  4452541000033114  4452641000033110  4549141000033118  4549241000033113  4945241000033117  4945341000033110  4957741000033118  5007641000033119  5007741000033111  5007841000033118  5007941000033114  5132241000033116  5576041000033115  5997041000033115  6029841000033115  6279741000033117  6391041000033117  7874941000033110  8115741000033119  8115841000033112  8115941000033116  8116041000033114  8242541000033112  8242641000033113  8242741000033116  8242841000033114  8348941000033113  8959841000033119  8959941000033110  9106041000033110  9106141000033114  9106241000033120  9106341000033112  9230641000033112  9230841000033114  9851541000033114  9851641000033110  9851741000033118  9851841000033112  10598841000033112  10598941000033116  10614141000033116  10614241000033112  10614341000033118  10614441000033112  10614541000033112  10614641000033114  10614741000033116  10614841000033110  11781341000033116  11781441000033112  12326541000033110  12326641000033112  12326741000033120  12593141000033116  12593241000033110  12593341000033116  12664441000033116  13429041000033112  13606641000033120  13803041000033110  13828541000033120 | Glucophage 500mg tablets  Glucophage 850mg tablets  Metformin 500mg tablets  Metformin 850mg tablets  Metformin 500mg/5ml oral suspension  Avandamet 1mg/500mg tablets  Avandamet 2mg/500mg tablets  Rosiglitazone 1mg / Metformin 500mg tablets  Rosiglitazone 2mg / Metformin 500mg tablets  Rosiglitazone 2mg / Metformin 1g tablets  Rosiglitazone 4mg / Metformin 1g tablets  Avandamet 2mg/1000mg tablets  Avandamet 4mg/1000mg tablets  Metformin 500mg modified-release tablets  Glucophage SR 500mg tablets  Metformin Hydrochloride Sugar free suspension 500 mg/5 ml  Metformin 500mg/5ml oral solution sugar free  Metsol 500mg/5ml oral solution  Pioglitazone 15mg / Metformin 850mg tablets  Competact 15mg/850mg tablets  Vildagliptin 50mg / Metformin 850mg tablets  Vildagliptin 50mg / Metformin 1g tablets  Eucreas 50mg/1000mg tablets  Eucreas 50mg/850mg tablets  Metformin 750mg modified-release tablets  Glucophage SR 750mg tablets  Metformin 1g modified-release tablets  Glucophage SR 1000mg tablets  Bolamyn SR 500mg tablets  Metformin 500mg oral powder sachets sugar free  Metformin 1g oral powder sachets sugar free  Glucophage 500mg oral powder sachets  Glucophage 1000mg oral powder sachets  Janumet 50mg/1000mg tablets  Metformin 1g / Sitagliptin 50mg tablets  Metformin 500mg/5ml oral solution  Metabet SR 500mg tablets  Metabet SR 1000mg tablets  Glucient SR 500mg tablets  Diagemet XL 500mg tablets  Linagliptin 2.5mg / Metformin 1g tablets  Linagliptin 2.5mg / Metformin 850mg tablets  Jentadueto 2.5mg/1000mg tablets  Jentadueto 2.5mg/850mg tablets  Saxagliptin 2.5mg / Metformin 850mg tablets  Saxagliptin 2.5mg / Metformin 1g tablets  Komboglyze 2.5mg/850mg tablets  Komboglyze 2.5mg/1000mg tablets  Bolamyn SR 1000mg tablets  Alogliptin 12.5mg / Metformin 1g tablets  Vipdomet 12.5mg/1000mg tablets  Dapagliflozin 5mg / Metformin 1g tablets  Dapagliflozin 5mg / Metformin 850mg tablets  Xigduo 5mg/1000mg tablets  Xigduo 5mg/850mg tablets  Sukkarto SR 500mg tablets  Sukkarto SR 1000mg tablets  Canagliflozin 50mg / Metformin 1g tablets  Canagliflozin 50mg / Metformin 850mg tablets  Vokanamet 50mg/850mg tablets  Vokanamet 50mg/1000mg tablets  Glucient SR 750mg tablets  Glucient SR 1000mg tablets  Empagliflozin 5mg / Metformin 850mg tablets  Empagliflozin 5mg / Metformin 1g tablets  Empagliflozin 12.5mg / Metformin 850mg tablets  Empagliflozin 12.5mg / Metformin 1g tablets  Synjardy 5mg/850mg tablets  Synjardy 5mg/1000mg tablets  Synjardy 12.5mg/850mg tablets  Synjardy 12.5mg/1000mg tablets  Metformin 1g/5ml oral solution sugar free  Metformin 850mg/5ml oral solution sugar free  Meijumet 500mg modified-release tablets  Meijumet 750mg modified-release tablets  Meijumet 1000mg modified-release tablets  Yaltormin SR 500mg tablets  Yaltormin SR 750mg tablets  Yaltormin SR 1000mg tablets  Metuxtan SR 500mg tablets  Sukkarto SR 750mg tablets  Glucorex SR 500mg tablets  Metformin 1g tablets  Axpinet 500mg tablets |

**CPRD GOLD**

| **Product Code** | **Product Name** |
| --- | --- |
| 23  93  735  2928  3252  6855  7048  7166  7325  7375  7610  7815  11601  11604  11609  11610  11717  11737  11760  11990  14164  16044  16213  17580  18220  20810  25678  26258  27501  30316  31077  31146  33087  33674  34004  34020  34135  34323  34504  34598  34697  34742  34836  34917  37874  37902  38355  38400  38551  39203  39560  39598  39729  39988  40007  40110  40233  42161  43270  43619  43684  44250  45581  46989  47939  48149  49502  49738  50570  50682  50821  50970  51080  51135  51527  52221  52442  52445  52449  52634  53478  53774  53867  54150  54442  54891  54898  54973  55270  55711  55739  56965  57147  57457  58051  58607  58865  59385  59620  60012  60074  60286  60497  60643  60968  61043  61559  62144  62265  62605  62824  63031  63045  63307  63929  64743  64939  65057  65059  65066  65083  65344  65694  65923  66008  66136  66854  66855  68203  68214  68389  68589  68636  69221  69370  70463  70477  70744  71012  71198  71890  72001  72046  72052  72107  72695  73252  73254  73285  73303  73460  73511  73525  73673  73808  73892  74021  74734  75700  75778  77753  78293  78392  78537  79017  80647  81428  81864  82085  82349  82372  82855  82890  83088  83608  83841  83988  84014  84627  84717  84743  84958  85047  85064  86710  87325  87341  87808  87891 | Metformin 500mg tablets  Metformin 850mg tablets  Metformin 100mg/ml Oral solution  METFORMIN HCl 850 MG TAB  METFORMIN HCl 500 MG TAB  Avandamet 2mg/500mg tablets (GlaxoSmithKline UK Ltd)  Metformin 500mg modified-release tablets  Glucophage 500mg tablets (Merck Serono Ltd)  Avandamet 4mg/1000mg tablets (GlaxoSmithKline UK Ltd)  Rosiglitazone 4mg / Metformin 1g tablets  Glucophage 850mg tablets (Merck Serono Ltd)  METFORMIN 800 MG TAB  Rosiglitazone 2mg / Metformin 500mg tablets  Rosiglitazone 1mg / Metformin 500mg tablets  Metformin with rosiglitazone 500mg + 1mg Tablet  Metformin with rosiglitazone 500mg + 2mg Tablet  Rosiglitazone 2mg / Metformin 1g tablets  Metformin with rosiglitazone 1000mg + 4mg Tablet  Metformin with rosiglitazone 1000mg + 2mg Tablet  Metformin 500mg/5ml oral solution sugar free  Avandamet 2mg/1000mg tablets (GlaxoSmithKline UK Ltd)  Glucophage SR 500mg tablets (Merck Serono Ltd)  METFORMIN 250 MG TAB  Avandamet 1mg/500mg tablets (GlaxoSmithKline UK Ltd)  Pioglitazone 15mg / Metformin 850mg tablets  METFORMIN  Glucamet 500mg Tablet (Opus Pharmaceuticals Ltd)  Glucamet 850mg Tablet (Opus Pharmaceuticals Ltd)  Orabet 500mg Tablet (Lagap)  Metformin with pioglitazone 850mg + 15mg Tablet  Competact 15mg/850mg tablets (Takeda UK Ltd)  Metsol 500mg/5ml oral solution (Kappin Ltd)  Metformin 500mg tablets (Actavis UK Ltd)  Metformin 850mg tablets (A A H Pharmaceuticals Ltd)  Metformin 500mg tablets (IVAX Pharmaceuticals UK Ltd)  Metformin 850mg tablets (IVAX Pharmaceuticals UK Ltd)  Metformin 500mg Tablet (M & A Pharmachem Ltd)  Metformin 500mg tablets (A A H Pharmaceuticals Ltd)  Metformin 500mg tablets (Wockhardt UK Ltd)  Metformin 500mg tablets (Mylan)  Metformin 850mg tablets (Wockhardt UK Ltd)  Metformin 850mg tablets (Teva UK Ltd)  Metformin 850mg tablets (Actavis UK Ltd)  Metformin 500mg tablets (Teva UK Ltd)  Vildagliptin 50mg / Metformin 850mg tablets  Vildagliptin 50mg / Metformin 1g tablets  Metformin 750mg modified-release tablets  Glucophage SR 750mg tablets (Merck Serono Ltd)  Eucreas 50mg/1000mg tablets (Novartis Pharmaceuticals UK Ltd)  Eucreas 50mg/850mg tablets (Novartis Pharmaceuticals UK Ltd)  Bolamyn SR 500mg tablets (Teva UK Ltd)  Metformin 1g modified-release tablets  Glucophage SR 1000mg tablets (Merck Serono Ltd)  Metformin 500mg oral powder sachets sugar free  Glucophage 1000mg oral powder sachets (Merck Serono Ltd)  Glucophage 500mg oral powder sachets (Merck Serono Ltd)  Metformin 1g oral powder sachets sugar free  Orabet 500mg Tablet (Sandoz Ltd)  Metformin 500mg/5ml oral solution sugar free (Rosemont Pharmaceuticals Ltd)  Metformin 1g / Sitagliptin 50mg tablets  Janumet 50mg/1000mg tablets (Merck Sharp & Dohme Ltd)  Metformin 500mg/5ml Oral solution (Hillcross Pharmaceuticals Ltd)  Metabet SR 500mg tablets (Morningside Healthcare Ltd)  Metabet SR 1000mg tablets (Morningside Healthcare Ltd)  Glucient SR 500mg tablets (Consilient Health Ltd)  Metformin 500mg tablets (Almus Pharmaceuticals Ltd)  Glucophage SR 500mg tablets (Mawdsley-Brooks & Company Ltd)  Metformin 1g modified-release tablets (A A H Pharmaceuticals Ltd)  Glucophage SR 500mg tablets (Lexon (UK) Ltd)  Jentadueto 2.5mg/1000mg tablets (Boehringer Ingelheim Ltd)  Metformin 850mg tablets (Pfizer Ltd)  Metformin 500mg tablets (Bristol Laboratories Ltd)  Metabet SR 1000mg tablets (Actavis UK Ltd)  Metformin 500mg modified-release tablets (A A H Pharmaceuticals Ltd)  Metformin 500mg tablets (Boston Healthcare Ltd)  Diagemet XL 500mg tablets (Thornton & Ross Ltd)  Metformin 500mg tablets (Pfizer Ltd)  Linagliptin 2.5mg / Metformin 1g tablets  Linagliptin 2.5mg / Metformin 850mg tablets  Glucophage SR 500mg tablets (DE Pharmaceuticals)  Metformin 500mg modified-release tablets (Kent Pharmaceuticals Ltd)  Metabet SR 500mg tablets (Actavis UK Ltd)  Metformin 500mg tablets (Zentiva)  Jentadueto 2.5mg/850mg tablets (Boehringer Ingelheim Ltd)  Metformin (roi) 1000mg Tablet  Saxagliptin 2.5mg / Metformin 1g tablets  Metformin 850mg tablets (Almus Pharmaceuticals Ltd)  Saxagliptin 2.5mg / Metformin 850mg tablets  Duformin 500mg Tablet (Dumex Ltd)  Metformin 500mg tablets (Alliance Healthcare (Distribution) Ltd)  Metformin 500mg tablets (Tillomed Laboratories Ltd)  Komboglyze 2.5mg/1000mg tablets (AstraZeneca UK Ltd)  Bolamyn SR 1000mg tablets (Teva UK Ltd)  Metformin 500mg tablets (Milpharm Ltd)  Metformin 500mg/5ml oral solution  Metformin 500mg/5ml oral solution sugar free (Zentiva)  Komboglyze 2.5mg/850mg tablets (AstraZeneca UK Ltd)  Vipdomet 12.5mg/1000mg tablets (Takeda UK Ltd)  Glucophage SR 500mg tablets (Waymade Healthcare Plc)  Dapagliflozin 5mg / Metformin 1g tablets  Metformin 1g modified-release tablets (Waymade Healthcare Plc)  Metformin 500mg/5ml oral suspension  Alogliptin 12.5mg / Metformin 1g tablets  Xigduo 5mg/1000mg tablets (AstraZeneca UK Ltd)  Metformin 500mg modified-release tablets (Actavis UK Ltd)  Sukkarto SR 1000mg tablets (Morningside Healthcare Ltd)  Sukkarto SR 500mg tablets (Morningside Healthcare Ltd)  Metformin 500mg modified-release tablets (DE Pharmaceuticals)  Metformin 500mg modified-release tablets (Mawdsley-Brooks & Company Ltd)  Metformin 850mg tablets (Kent Pharmaceuticals Ltd)  Metformin 1g modified-release tablets (Actavis UK Ltd)  Dapagliflozin 5mg / Metformin 850mg tablets  Metformin 850mg tablets (Relonchem Ltd)  Metformin 1g/5ml oral solution  Canagliflozin 50mg / Metformin 1g tablets  Canagliflozin 50mg / Metformin 850mg tablets  Glucient SR 1000mg tablets (Consilient Health Ltd)  Empagliflozin 5mg / Metformin 1g tablets  Xigduo 5mg/850mg tablets (AstraZeneca UK Ltd)  Empagliflozin 12.5mg / Metformin 1g tablets  Synjardy 5mg/1000mg tablets (Boehringer Ingelheim Ltd)  Empagliflozin 5mg / Metformin 850mg tablets  Metformin 500mg modified-release tablets (Waymade Healthcare Plc)  Metformin 1g modified-release tablets (Mawdsley-Brooks & Company Ltd)  Synjardy 12.5mg/1000mg tablets (Boehringer Ingelheim Ltd)  Glucophage SR 1000mg tablets (Waymade Healthcare Plc)  Vokanamet 50mg/1000mg tablets (Napp Pharmaceuticals Ltd)  Empagliflozin 12.5mg / Metformin 850mg tablets  Metformin 500mg modified-release tablets (Almus Pharmaceuticals Ltd)  Metformin 500mg/5ml oral solution sugar free (A A H Pharmaceuticals Ltd)  Metformin 500mg/5ml oral solution sugar free (Pinewood Healthcare)  Metformin 1g/5ml oral solution sugar free  Metformin 850mg/5ml oral solution sugar free  Metformin 1g oral powder sachets sugar free (J M McGill Ltd)  Synjardy 5mg/850mg tablets (Boehringer Ingelheim Ltd)  Synjardy 12.5mg/850mg tablets (Boehringer Ingelheim Ltd)  Glucient SR 750mg tablets (Consilient Health Ltd)  Metformin 500mg/5ml oral solution sugar free (Focus Pharmaceuticals Ltd)  Metformin 500mg tablets (Phoenix Healthcare Distribution Ltd)  Metformin 500mg tablets (Zanza Laboratories Ltd)  Metformin 500mg/5ml oral solution sugar free (Almus Pharmaceuticals Ltd)  Metformin 1g modified-release tablets (DE Pharmaceuticals)  Metformin 500mg tablets (DE Pharmaceuticals)  Metformin 500mg Tablet (Lagap)  Meijumet 500mg modified-release tablets (Medreich Plc)  Metformin 500mg Tablet (Celltech Pharma Europe Ltd)  Yaltormin SR 500mg tablets (Wockhardt UK Ltd)  Yaltormin SR 1000mg tablets (Wockhardt UK Ltd)  Metformin 500mg tablets (Waymade Healthcare Plc)  Metformin 500mg/5ml oral solution sugar free (Colonis Pharma Ltd)  Metformin 500mg/5ml oral solution sugar free (Actavis UK Ltd)  Yaltormin SR 750mg tablets (Wockhardt UK Ltd)  Metformin 500mg/5ml oral solution sugar free (Alliance Healthcare (Distribution) Ltd)  Metformin 500mg/5ml oral solution sugar free (Sigma Pharmaceuticals Plc)  Metformin 500mg/5ml oral solution sugar free (Actavis UK Ltd)  Metformin 500mg tablets (Relonchem Ltd)  Metformin 250mg/5ml oral solution  Metformin 850mg capsules  Metformin 500mg/5ml oral solution sugar free (Waymade Healthcare Plc)  Metformin 500mg tablets (Crescent Pharma Ltd)  Meijumet 1000mg modified-release tablets (Medreich Plc)  Alogliptin 12.5mg / Metformin 1g tablets (Colorama Pharmaceuticals Ltd)  Metformin 850mg tablets (Zentiva)  Metformin 850mg/5ml oral suspension  Metformin 850mg tablets (Mylan)  Metuxtan SR 500mg tablets (Accord Healthcare Ltd)  Sukkarto SR 750mg tablets (Morningside Healthcare Ltd)  Metformin 500mg modified-release tablets (Morningside Healthcare Ltd)  Metformin 500mg tablets (RX Farma)  Metformin 850mg tablets (Phoenix Healthcare Distribution Ltd)  Competact 15mg/850mg tablets (DE Pharmaceuticals)  Metformin 500mg modified-release tablets (Medihealth (Northern) Ltd)  Metformin 850mg tablets (Milpharm Ltd)  Glucorex SR 500mg tablets (GlucoRx Ltd)  Metformin 500mg modified-release tablets (Consilient Health Ltd)  Metformin 500mg tablets (Sigma Pharmaceuticals Plc)  Metformin 1g modified-release tablets (Morningside Healthcare Ltd)  Metformin 1g tablets  Metformin 1g modified-release tablets (Accord Healthcare Ltd)  Metformin 500mg Tablet (Ratiopharm UK Ltd)  Metformin Oral solution  Pioglitazone 15mg / Metformin 850mg tablets (A A H Pharmaceuticals Ltd)  Metformin 1g modified-release tablets (Medihealth (Northern) Ltd)  Metformin 500mg modified-release tablets (A A H Pharmaceuticals Ltd)  Metformin 750mg modified-release tablets (DE Pharmaceuticals)  Glucophage SR 1000mg tablets (Pilsco Ltd)  Metformin 1g/5ml oral solution sugar free (Colonis Pharma Ltd)  Metformin 1g modified-release tablets (A A H Pharmaceuticals Ltd)  Metformin 500mg modified-release tablets (Amarox Ltd) |

Supplemental Table 2. Characteristics of patients prescribed a second-generation antipsychotic and metformin (≤1 month before to ≤2 years after SGA prescription) with a diagnosis of diabetes and/or PCOS compared to those without a recorded diagnosis of diabetes or PCOS.

| **Characteristic** | **Metformin  (no diabetes or PCOS) N = 341** | | **Metformin  (with diagnosis and/or PCOS)  N = 355** |
| --- | --- | --- | --- |
| **Sex, N (%)** |  | |  |
| Female | 186 (54.55%) | | 186 (52.39%)/ |
| Male | 155 (45.45%) | | 169 (47.61%) |
| **Age, Mean (SD)** |  | |  |
| at index date | 52.32 (14.33) | | 54.77 (14.15) |
| at first SGA prescription | 49.24 (14.12) | | 51.74 (14.45) |
| at first SMI diagnosis | 44.55 (15.47) | | 45.24 (16.03) |
| **Ethnicity, N (%)** |  | |  |
| Asian | 40 (12.58%) | | 37 (11.42%) |
| Black | 36 (11.32%) | | 29 (8.95%) |
| Mixed/Other | 11 (3.46%) | | 6 (1.86%) |
| White | 231 (72.64%) | | 252 (77.78%) |
| *Unknown* | *23* | | *31* |
| **SGA initiated at index date, N (%)** |  | |  |
| Aripiprazole | 52 (15.25%) | | 54 (15.21%) |
| Olanzapine | 125 (36.66%) | | 106 (29.86%) |
| Risperidone | 74 (21.7%) | | 94 (26.48%) |
| Quetiapine | 90 (26.39%) | | 101 (28.45%) |
| **SMI diagnosis, N (%)** |  | |  |
| Bipolar disorder | 141 (41.35%) | | 143 (40.28%) |
| Other non-organic psychoses | 126 (36.95%) | | 110 (30.99%) |
| Schizophrenia | 74 (21.70%) | | 102 (28.73%) |
| **Geographical region, N (%)** |  | |  |
| East Midlands | 8 (2.35%) | | 5 (1.41%) |
| East Of England | 14 (4.11%) | | 12 (3.38%) |
| London | 87 (25.51%) | | 75 (21.13%) |
| North East | 11 (3.23%) | | 7 (1.97%) |
| North West | 66 (19.35%) | | 56 (15.77%) |
| Northern Ireland | 4 (1.17%) | | 6 (1.69%) |
| Scotland | 17 (4.99%) | | 23 (6.48%) |
| South East | 45 (13.20%) | | 49 (13.80%) |
| South West | 24 (7.04%) | | 35 (9.86%) |
| Wales | 21 (6.16%) | | 23 (6.48%) |
| West Midlands | 36 (10.56%) | | 52 (14.65%) |
| Yorkshire & The Humber | 8 (2.35%) | | 12 (3.38%) |
| **2019 English IMD quintile, N (%)** | |  | |
| 1 (Least Deprived) | 24 (8.14%) | | 33 (11.19%) |
| 5 (Most Deprived) | 107 (36.27%) | | 96 (32.54%) |
| *Unknown* | *46* | | *60* |
| **Baseline Weight (kg), Mean (SD)** | 92.78 (24.53) | | 88.42 (22.42) |
| **Baseline BMI, N (%)** |  | |  |
| Underweight | 1 (0.39%) | | 6 (1.89%) |
| Healthy | 29 (11.24%) | | 50 (15.72%) |
| Obese | 59 (22.87%) | | 87 (27.36%) |
| Overweight | 169 (65.50%) | | 175 (55.03%) |
| *Unknown* | *83* | | *37* |
| **Comorbidities, N (%)** |  | |  |
| Cerebrovascular disease | 16 (4.69%) | | 27 (7.61%) |
| Myocardial infarction | 16 (4.69%) | | 11 (3.10%) |
| Liver disease | 11 (3.23%) | | 6 (1.69%) |
| Renal disease | 19 (5.57%) | | 29 (8.17%) |
| Hypertension | 122 (35.78%) | | 151 (42.54%) |
| Dyslipidaemia | 79 (23.17%) | | 90 (25.35%) |
| **Other medications prescribed in prior 2y, N (%)** | | | |
| Antidepressants | 224 (65.69%) | | 203 (57.18%) |
| Lipid-regulating medications | 104 (30.50%) | | 189 (53.24%) |
| Insulin | 0 (0.00%) | | 25 (7.04%) |
| **Prior Exposure to Substances, N (%)** | | | |
| Alcohol misuse | 31 (9.09%) | | 34 (9.58%) |
| Substance misuse | 33 (9.68%) | | 18 (5.07%) |
| Ex-Smoker | 52 (15.34%) | | 52 (14.73%) |
| Current Smoker | 159 (46.90%) | | 155 (43.91%) |
| **Biochemical parameters,^1^ Mean (SD)** | |  | |
| HbA1c | 47.09 (14.34) | | 57.63 (21.14) |
| *Unknown* | *219* | | *45* |
| Glucose | 6.41 (2.26) | | 8.84 (4.42) |
| *Unknown* | *46* | | *66* |

SGA, second generation antipsychotic; IMD, index of multiple deprivation; BMI, body mass index; HbA1c, glycated hemoglobin.

Percentages are of patients with non-missing data.

^1^ Biochemical parameters are defined according to test result values recorded on or within the two years prior to the index date, using the value closest to the index date. For values requiring a blood test for measurement, values recorded up to seven days after the index date were also considered on the assumption that results might relate to the date on which test results were received, rather than the date on which the blood test was taken.

Supplemental Table 3. Mean absolute weight and percentage change over time in patients prescribed SGA only versus those prescribed SGA+Metformin (initiated ≤1 month before or ≤3 months after SGA prescription).

| **Time** | **SGA Only (N=21,885)** | | **SGA+Metformin  (N=212)** | |
| --- | --- | --- | --- | --- |
|  | *Body weight (kg)* | *Change (%)* | *Body weight (kg)* | *Change (%)* |
| Baseline | 76.06 (75.75, 76.37) | - | 90.44 (86.72, 94.17) | - |
| 6m | 78.10 (77.61, 78.59) | 3.18 (2.15, 4.20) | 89.86 (86.02, 93.69) | -0.20 (-3.90, 3.49) |
| 1y | 78.48 (77.53, 79.44) | 3.80 (1.28, 6.31) | 89.54 (85.74, 93.35) | -0.44 (-4.18, 3.29) |
| 2y | 78.71 (75.80, 81.61) | 4.16 (-1.26, 9.58) | 89.33 (85.16, 93.50) | -0.65 (-4.26, 2.96) |

Values are mean with 95% confidence intervals. Missing data was imputed using multiple imputation. Estimates were pooled according to Rubin's rules. Change is the percentage change from baseline.
